# A Dynamic Bayesian Model for Identifying High-Mortality Risk in Hospitalized COVID-19 Patients

**DOI:** 10.1101/2021.02.02.21251023

**Authors:** Amir Momeni-Boroujeni, Rachelle Mendoza, Isaac J. Stopard, Ben Lambert, Alejandro Zuretti

## Abstract

**Introduction:** As COVID-19 hospitalization rates remain high, there is an urgent need to identify prognostic factors to improve treatment. Our analysis, to our knowledge, is one of the first to quantify the risk associated with dynamic clinical measurements taken throughout the course of hospitalization.

**Methods:** We collected data for 553 PCR-positive COVID-19 patients admitted to hospital whose eventual outcomes were known. The data collected for the patients included demographics, comorbidities and laboratory values taken at admission and throughout the course of hospitalization. We trained multivariate Markov prognostic models to identify high-risk patients at admission along with a dynamic measure of risk incorporating time-dependent changes in patients’ laboratory values.

**Results:** From the set of factors available upon admission, the Markov model determined that age >80 years, history of coronary artery disease and chronic obstructive pulmonary disease increased mortality risk. The lab values upon admission most associated with mortality included neutrophil percentage, RBC, RDW, protein levels, platelets count, albumin levels and MCHC. Incorporating dynamic changes in lab values throughout hospitalization lead to dramatic gains in the predictive accuracy of the model and indicated a catalogue of variables for determining high-risk patients including eosinophil percentage, WBC, platelets, pCO2, RDW, LUC count, alkaline phosphatase and albumin.

**Conclusion:** Our prognostic model highlights the nuance of determining risk for COVID-19 patients and indicates that, rather than a single variable, a range of factors (at different points in hospitalization) are needed for effective risk stratification.

## INTRODUCTION

As global COVID-19 deaths exceed one million (1), improved clinical care remains essential to reducing the health impact of the pandemic. Severe SARS-CoV-2 infection often manifests as pneumonia and may cause multiple organ damage, including acute respiratory distress syndrome (2), acute kidney injury (2, 3), acute myocardial injury (4), hepatic injury (2) and coagulopathy (5). Abnormalities in the surrogate laboratory test corresponding to these organ damages have been reported in COVID-19 patients including elevated troponin (6, 7) and D-dimer (5), hypoxemia, hypercapnia (8), thrombocytopenia (9) and prolonged prothrombin time. The latter were also found to be of prognostic significance along with bilirubin, blood urea and albumin levels (10). Elevated C-reactive protein (CRP), ferritin, lactate dehydrogenase (LDH) and procalcitonin, and lymphopenia, are also associated with mortality (11). In addition, the risk of adverse outcomes has been associated with patient characteristics such as advanced age (12), male sex (13) and existing comorbidities (14).

The identification of prognostic factors and the development of prognostic models, which aim to predict the course of infection of hospitalized patients, are necessary to inform clinical decisions and guide patient care (15, 16). Efficient COVID-19 transmission, a high infection fatality ratio (17) and underprepared health systems (18) have seen many hospitals exceed capacity (19, 20). In this context, accurate prognostication, alongside ethical considerations (21, 22), is critical for patient triage and healthcare resource management (16).

By combining multiple variables into a single analysis, prognostic models can be used to investigate the relative importance of different prognostic factors and evaluate their impact on mortality risk (23). Since the beginning of the outbreak, numerous prognostic models, applying regression, survival analysis or decision trees, have been developed. For applicability, these models typically consider a small number of variables and an outcome at a single follow up time (16, 24). A number of studies have demonstrated that certain laboratory values for COVID-19 patients change through disease progression, however, to our knowledge, none have used these to quantify a dynamic measure of mortality risk (25-27).

Our aim was to develop a prognostic model for hospitalized COVID-19 patients which incorporates dynamic laboratory value data along with patients’ admission profiles: allowing us to identify key determinants of risk.

## Methods

### Case Selection and Data Extraction

Approval for the study was obtained from the State University of New York, Downstate Medical Center Institutional Review Board (IRB#1595271-1).

A retrospective query was performed among patients admitted to SUNY Downstate Medical Center with COVID-19-related symptoms and confirmed PCR-positive from early February 2020 until the end of March 2020. Stratified randomization was used to select at least 200 patients who were discharged and 200 patients who died due to COVID-19 complications. Patients whose outcome was unknown were excluded. The outcome for patients was recorded as either “discharged” or “COVID-19 related mortality”. Demographic, clinical history and laboratory data were extracted from the hospital’s electronic health records.

### Models of dynamic risk

The data were processed to convert them into a form amenable to estimation (see supplementary materials (SOM)), resulting in a few individuals and tests being dropped from the analysis (mainly due to missing data). This meant that 475 individuals and 28 laboratory tests were included in our models.

Two sets of analyses were conducted: the first was a logistic regression analysis, which aimed to determine those factors most predictive of patient mortality. The second was a Markov model, which analyzed the dynamic sequence of observations for each patient throughout their stay and aimed to examine how changes in these variables affected the probability a patient was discharged or died on a given day (Figure 1; SOM). Both models were estimated in a Bayesian framework (details in SOM) and, as such, there is no need for an arbitrary cutoff representing whether a factor is significant: any probabilities reported represent the posterior probability that a given variable had an odds ratio exceeding one.

**Figure 1.**
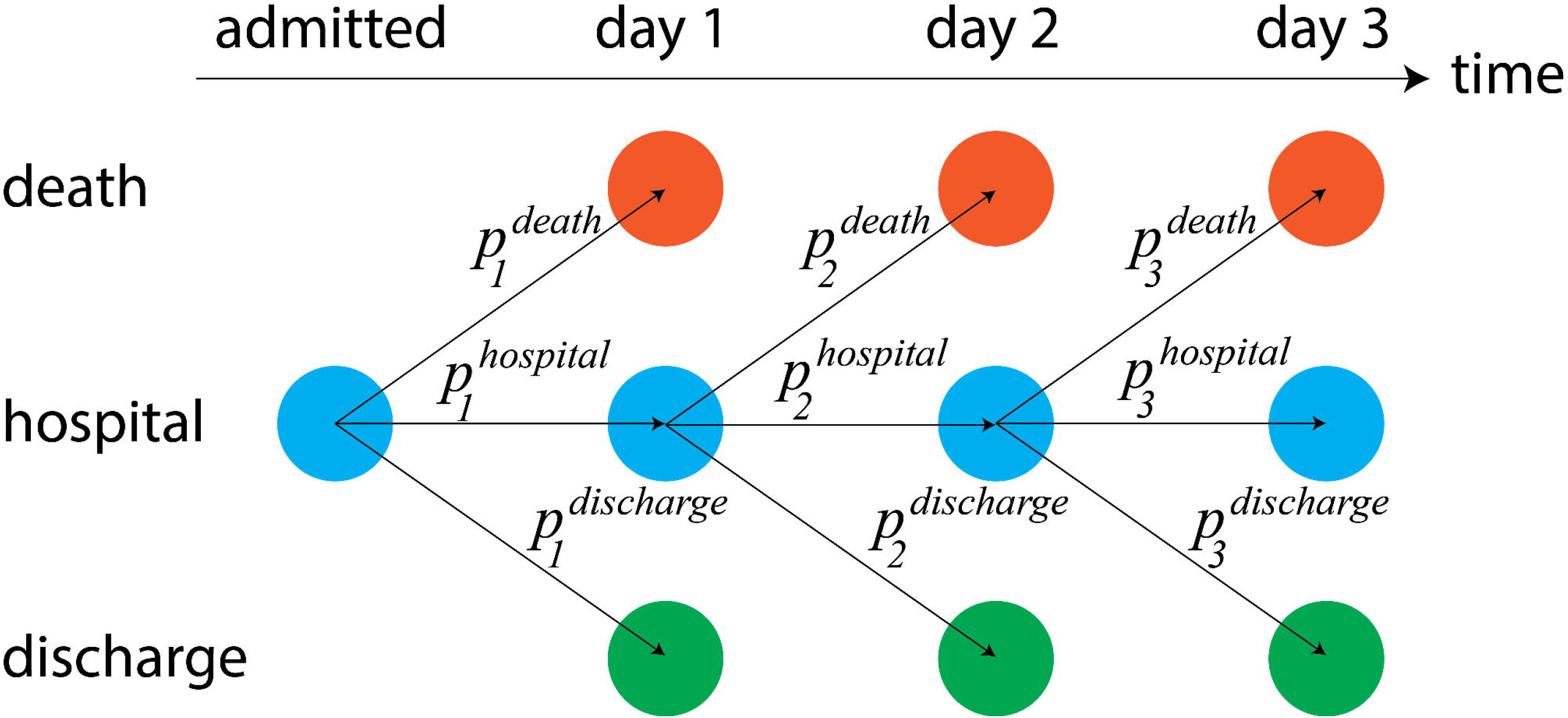
Markov model schematic: Colors indicate each of the three distinct states. Arrows indicate potential changes in the patient’s state from one day to the next. These state transitions are influenced by probabilities which, in the model, can vary with patient characteristics, timing of their admission, initial lab values and dynamic changes in values.

For both approaches, we estimated a series of models including different sets of predictors. The first set (“patient” variables) included demographic characteristics. The second set (“pat. + comorbidities”) supplemented the background variables with patient comorbidities. The third set (“admission”) supplemented the previous with the initial measurements for the laboratory tests. The final regression (“post-admission”) then included percentage changes in each laboratory value from that at admission.

## Results

### Patient Characteristics

We collected data for 553 patients. Here, we describe the patient characteristics prior to data processing required for estimation.

The cohort consisted of 342 discharged patients and 211 expired patients. There were 271 (50.3%) females and 268 (49.7%) males. The median age was 69 years (range: 6-101 years). Most of the patients hospitalized were black (n=472, 86.8%; Table 1).

**Table 1:** Summary of patient characteristics. Note, that a number of variables have missing observations meaning that the totals across all groups do not sum to the total number of patients (n=553): for example, there were 14 patients whose sex was not recorded.

| Category | Variable | Count | Frequency |
| --- | --- | --- | --- |
| Outcome | Outcome: Discharged | 342 | 61.8% |
|  | Outcome: Expired | 211 | 38.2% |
| Demographic | Sex: Female | 271 | 50.3% |
|  | Sex: Male | 268 | 49.7% |
|  | Age: 0-40 | 26 | 4.8% |
|  | Age: 40-50 | 43 | 7.9% |
|  | Age: 50-60 | 93 | 17.2% |
|  | Age: 60-70 | 140 | 25.8% |
|  | Age: 70-80 | 137 | 25.3% |
|  | Age: 80-100 | 103 | 19.0% |
|  | Ethnicity: Black | 472 | 86.8% |
|  | Ethnicity: Caucasian | 25 | 4.6% |
|  | Ethnicity: Hispanic | 17 | 3.1% |
|  | Ethnicity: Other or Unrecorded | 39 | 7.1% |
| Comorbidities | Asthma | 24 | 4.4% |
|  | Cancer | 16 | 2.9% |
|  | Cerebrovascular Disease | 25 | 4.6% |
|  | Congestive Heart Failure | 23 | 4.2% |
|  | Chronic Kidney Disease | 19 | 3.5% |
|  | Chronic Obstructive Pulmonary Disease | 25 | 4.6% |
|  | Coronary Artery Disease | 44 | 8.0% |
|  | Dementia | 13 | 2.4% |
|  | Diabetes | 229 | 41.9% |
|  | End-Stage Renal Disease | 54 | 9.9% |
|  | Hepatitis | 4 | 0.7% |
|  | Hyperlipidemia | 103 | 18.8% |
|  | Hypertension | 350 | 64.0% |

The most common comorbidity was hypertension (n= 350, 64.0%) followed by diabetes (n=229, 41.9%), hyperlipidemia (n= 103, 18.8%), end-stage renal disease (n=54, 9.9%) and coronary artery disease (CAD) (n=44, 8.0%). There was considerable within-patient clustering of the comorbidities (Figure S1).

We defined the test values at admission as the mean of those taken during the first day of hospitalization (Table S1). At admission, most patients had marked increase in CRP (median: 149 mg/L, Interquartile range (IQR): 80-246 mg/L), LDH (median: 468 IU/L, IQR: 342-638 IU/L) and ferritin (median: 879 ng/mL, IQR: 415-2132 ng/mL) levels. The patients also had decreased lymphocyte percentage (median: 12%, IQR: 8%-16%). Patients also tended to have increased blood urea nitrogen (BUN; median: 27 mg/dL, IQR: 16-50 mg/dL).

### Characteristics of discharged and expired patients

Expired patients were generally older (median: 73 versus 65 years old; *t*540 = 7.4, p < 0.01), more likely to be male (57% versus 45%: posterior overlap p < 0.01) and to have hypertension, diabetes, hyperlipidemia, CAD, cerebrovascular disease, cancer or dementia (Figure 2A).

**Figure 2.**
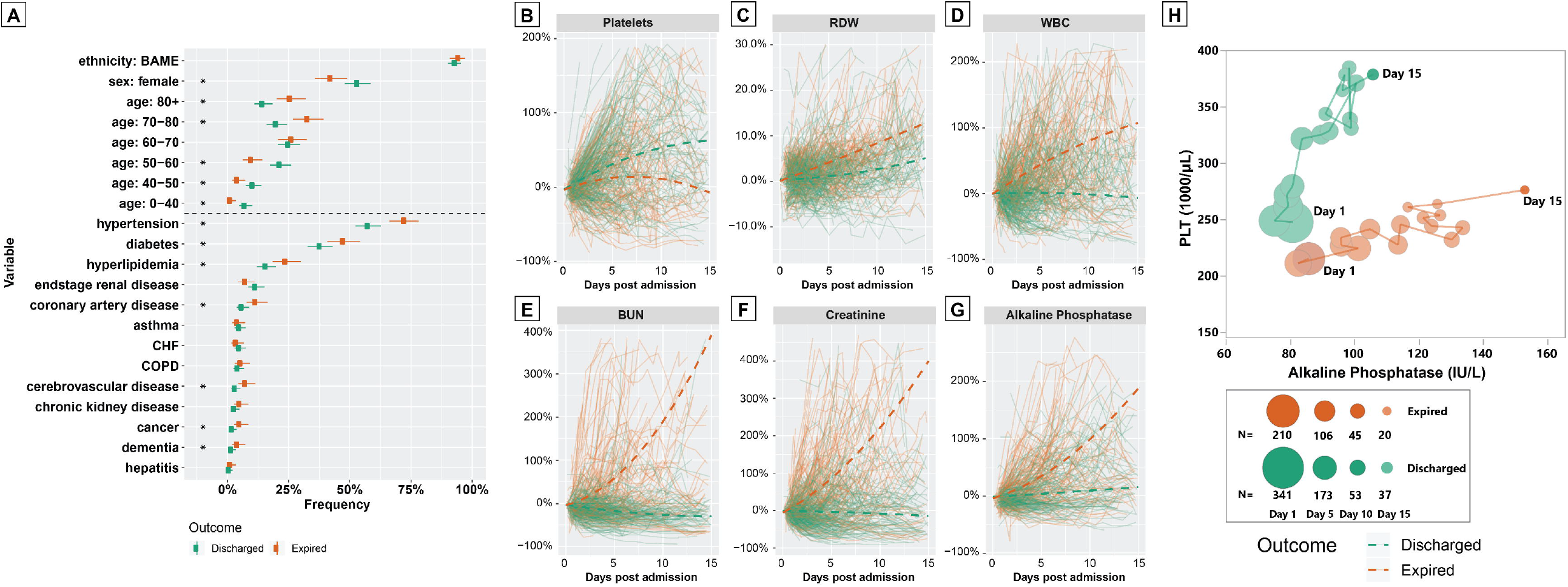
(A) Characteristics of patients according to outcome: The points indicate posterior median frequencies and the upper and lower whiskers indicate 2.5% and 97.5% posterior quantiles, calculated assuming a uniform prior and binomial likelihood. Asterisks indicate that the overlap in posteriors for each outcome was less than 0.05 for a given variable. (B-H) Time-series graphs of select laboratory values in hospitalized COVID-19 patients: Panels B-G show individual patient trajectories (solid lines) colored according to outcome for a selection of tests; dashed lines show fitted regression lines (see SOM) for patients grouped by outcome). Panel H shows daily averages of platelets count (vertical axis) versus alkaline phosphatase levels (horizontal axis) for all patients remaining in hospital during the first 15 days of hospitalization. Each point indicates the paired mean of both tests for one of the two patient groups.

For 27 of 38 tests considered, there were significant differences in the initial test values between the discharged and the expired patients (Table S2). Notably, expired patients had higher CRP (median: 192 versus 124 mg/L) and LDH levels (median: 551 versus 427 IU/L), and lower lymphocyte percentage (median: 10.4% versus 12.8%), platelets (median: 188 versus 223 10^3/µL), and albumin levels (median: 3.3 versus 3.5 g/dL).

We then compared the trends in test values throughout patients’ hospitalization course and analyzed the trends in test values as percentage changes from the values upon admission. To do so, we determined trends for patients grouped by outcome separately using linear mixed effects models (see SOM). Most tests showed visually distinct time trends for the two groups, indicating that, throughout hospitalization, those who went onto survive diverged from those who would not (Figure 2B-G, Figure S3). For example, patients who expired displayed less marked increases in platelet levels over time (Fig. 2B) but exhibited relative increases in red cell distribution width (RDW; Fig. 2C), white blood cell (WBC; Fig. 2D), BUN (Fig. 2E), creatinine (Fig. 2F) and alkaline phosphatase (ALKP; Fig 2G) levels. Plotting the average tests scores for platelets and ALKP over time shows that it is possible to further separate survivors from those who died (Fig. 2G). Indeed, this principle underpins the multivariate approach to determining risk factors in our dynamic Markov model.

### Analysis of dynamic patient risk

#### Univariate analysis

We used univariate survival analyses to illustrate the baseline patient characteristics and lab values upon admission that, individually, were the strongest determinants of risk. Increased age was the only statistically significant demographic risk factor (Table S3; Fig. S2). From the comorbidities, only hyperlipidemia and CAD led to increases in mortality risk. Of the initial lab values, 17 were associated with mortality: the tests whose high values were most associated with mortality were total bilirubin (OR: 1.25), followed by mean platelet volume (OR: 1.15). Conversely, increased eosinophil percentage (OR: 0.67), large unstained cells percentage (OR: 0.81) and platelets level (OR: 0.99) were associated with lower mortality.

### Multivariate analysis

We used our Markov model with different combinations of variables to determine patient risk at admission and throughout the course of hospitalization. The multivariate results indicate fewer variables had a strong impact on risk of death; by controlling for more factors, certain variables, for example, age and sex, become less important predictors (Fig. 3; Fig. S4). Contrastingly, some factors, like CAD and chronic obstructive pulmonary disease (COPD), became more important. Similar results were also obtained by logistic regression analysis (Table S4), which considered only patients’ outcomes: not the time taken for the outcome to occur. These results suggest that patient prognosis should be based on multiple factors rather than individual variables such as age.

**Figure 3.**
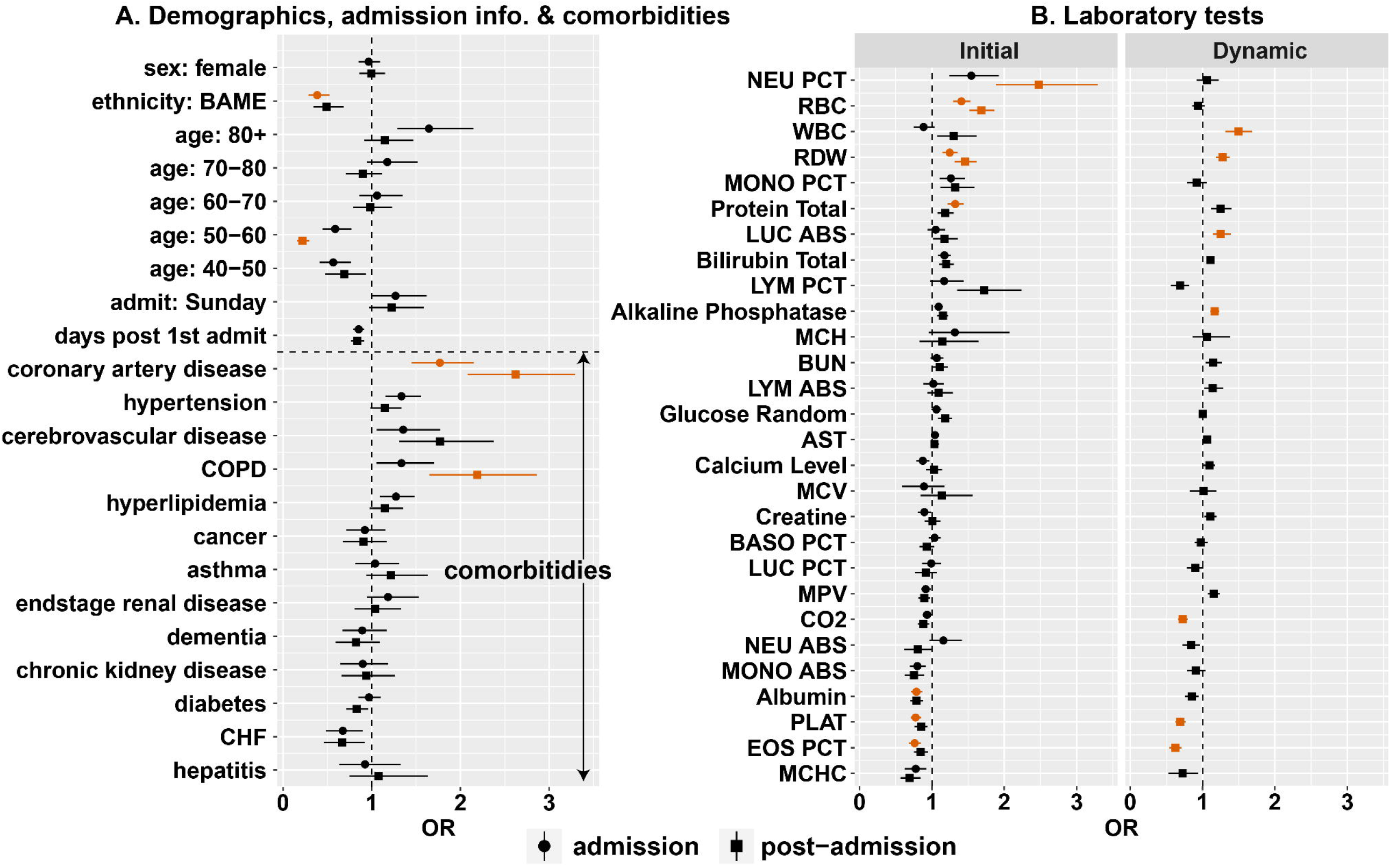
Estimated odds ratios for daily risk of death from Markov model: Panel A shows odds ratios (ORs) for demographic factors, variables associated with admission timing and comorbidities; Panel B shows ORs associated with lab values: both those upon admission (“Initial”) and % changes in lab values from these initial values (“Dynamic”). Marker types indicates the selection of regressors included in the model (see main text). Orange points indicate those odds ratios where the 5%-95% posterior quantiles did not cross zero. Note, that the results for the lab values were obtained on data that had been standardized so that differences best reflect clinically relevant differences (see main text).

Controlling for all the variables available upon admission, individuals aged over 80 were at higher risk (Fig. 3A, circular markers; Table S5; OR: 1.64, *Pr*(*OR* > 1) = 0.92). Upon admission, CAD (OR: 1.77, *Pr*(*OR* > 1) = 0.98), cerebrovascular disease (OR: 1.35, *Pr*(*OR* > 1) = 0.80) and COPD (OR: 1.33, *Pr*(*OR* > 1) = 0.81) were the comorbidities most associated with elevated mortality risk.

To ensure that our odds ratios across different lab tests upon admission were on comparable scale, we standardized data for each test meaning that odds ratios (shown in Fig. 3B) are associated with the degree to which a value is above the mean and relative to the standard deviation (Table S6). Consistent with previous studies, upon admission, high values of certain tests were associated with a lower risk of death including albumin (>3.4 g/dL, OR: 0.79, *Pr*(*OR* > 1) = 0.04); platelets (>236,000/μL, OR: 0.78, *Pr*(*OR* > 1) = 0.03); eosinophil percentage (>0.64%, OR: 0.76, *Pr*(*OR* > 1) = 0.03) and mean corpuscular haemoglobin concentration (MCHC; >30.9%, OR: 0.78, Pr(OR > 1) = 0.15). For other variables, high values were associated with increased risk: neutrophil percentage (>79.4%, OR: 1.55, *Pr*(*OR* > 1) = 0.92); red blood cell count (RBC; >4.47 x 106 cells/μL, OR: 1.41, *Pr*(*OR* > 1) = 1.00); monocyte percentage (MONO PCT; >5.0%, OR: 1.26, *Pr*(*OR* > 1) = 0.89); red cell distribution width (RDW; >14.9%, OR: 1.25, *Pr*(*OR* > 1) = 0.96) and total bilirubin (>0.74 mg/dL, OR: 1.17, Pr(OR > 1) = 0.92).

After a patient has been admitted, the predictive factors change as the results of ongoing laboratory tests are included (Figure 3, square markers). At this time, many of the variables most important at admission become less important predictors, for example, being older than 80 years is less predictive (OR: 1.15, *Pr*(*OR* > 1) = 0.66). Other variables, however, became more important predictors including CAD (OR: 2.62, Pr(OR > 1) = 1.00) and COPD (OR: 2.19, *Pr*(*OR* > 1) = 0.97). Many of the lab tests found important at admission continued to be so: high values of neutrophil percentage (OR: 2.48, *Pr*(*OR* > 1) = 0.99), RBC (OR: 1.68, *Pr*(*OR* > 1) = 1.00) and RDW (OR: 1.46, *Pr*(*OR* > 1) = 0.99) were associated with inflated mortality risk. Some of the admission values became more predictive, for example, high lymphocyte percentage was associated with higher risk (if >12.7%, OR: 1.72, *Pr*(*OR* > 1) = 0.94). Additionally, in the dynamic analysis, the risk of mortality increased the longer a patient was hospitalized (OR: 1.36, *Pr*(*OR* > 1) = 0.97).

In our analysis, we tracked the percentage change in each lab value relative to that at admission for each patient. Since different tests exhibited different dynamics over time (Fig. S2), we standardized these percentage changes so that the odds ratios estimated across the different tests were on a comparable scale (Table S6). For a number of tests, above average increases in values over time raised the chance of death including: white blood cell count (OR: 1.5, *Pr*(*OR* > 1) = 0.99), RDW (OR: 1.28, *Pr*(*OR* > 1) = 0.99), total protein (OR: 1.25, *Pr*(*OR* > 1) = 0.92), the absolute count of large unstained cells (LUC ABS; OR: 1.25, *Pr*(*OR* > 1) = 0.95), bilirubin total (OR: 1.11, *Pr*(*OR* > 1) = 0.95) and ALKP (OR: 1.17, *Pr*(*OR* > 1) = 0.97). Above average changes in other variables signaled lower risk including: EOS PCT (OR: 0.62, *Pr*(*OR* > 1) = 0.00), platelets (OR: 0.69, *Pr*(*OR* > 1) = 0.01), and CO2 (OR: 0.73, *Pr*(*O*R > 1) = 0.01).

To assess the internal validity of the Markov model (28), we performed k-fold cross-validation (see SOM). We performed this analysis for each of the four regressions performed. The first three regressions offered similar levels of mean accuracy: the *patient* regression was able to determine the patient outcome with an accuracy of 67% (Fig. S5); the *pat. + comorbidities* regression had an accuracy of 64%; and the *admission* regression had an accuracy of 67%. Including dynamic test values boosted predictive power to an accuracy of 83%.

### Dynamic measure of risk

We determined a dynamic measure of patient risk throughout hospitalization based on the Markov model which incorporated dynamic laboratory measurements. The model identified differences between the two groups upon admission: the group who went on to eventually be discharged had a mean probability of death on the first day of 0.015, whereas the expired group had a corresponding value more than four times greater. Throughout the course of hospitalization, the model could more accurately differentiate these groups, with the regression line for the discharged group remaining flat and that for the expired group increasing (Fig. 4A)—highlighting the importance of accounting for dynamic test values. Despite this, it was not possible to perfectly predict patient outcomes due to the strong overlap in estimated probabilities between the groups (see selected individuals in Figure 4A).

**Figure 4.**
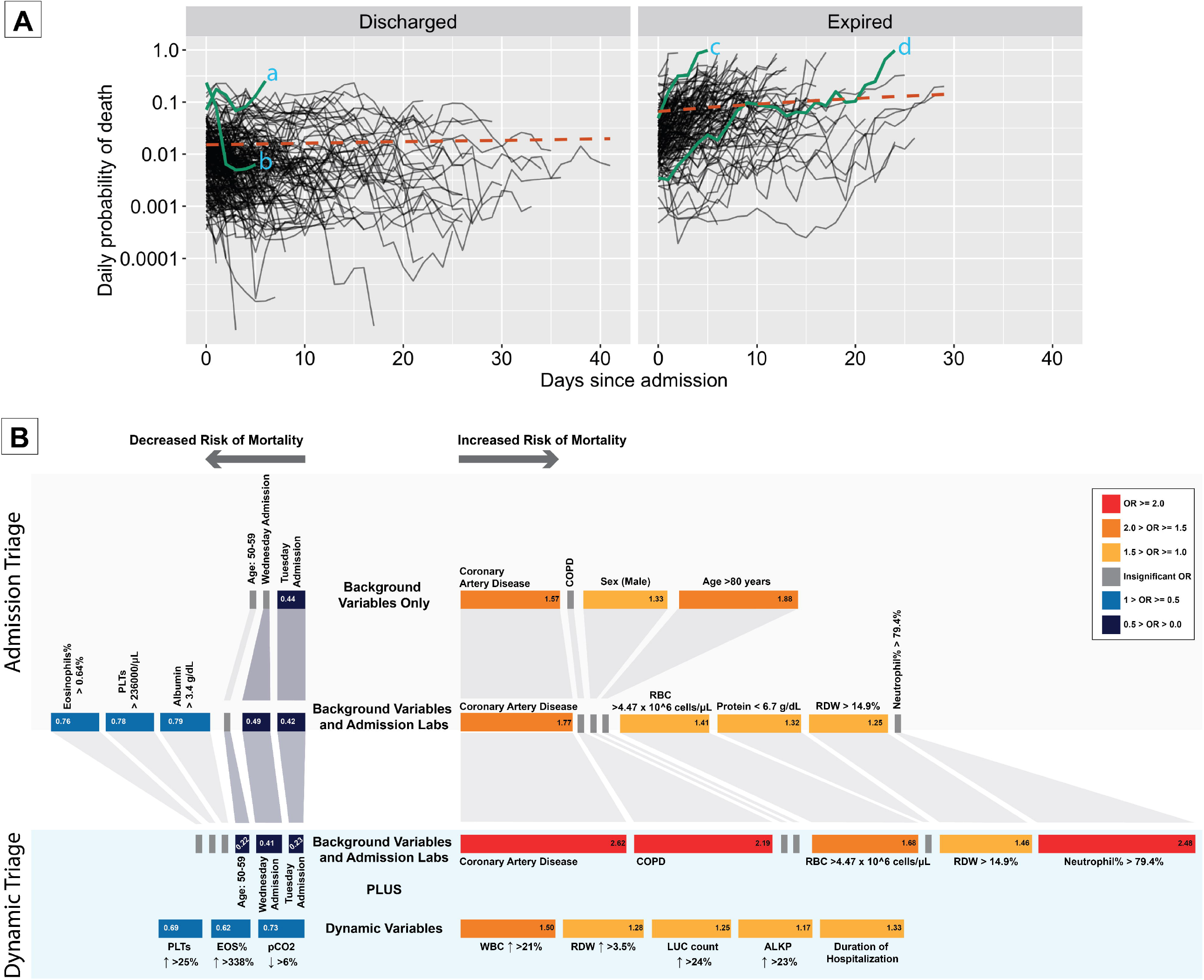
(A) Dynamic risk estimates for individual patients: Solid lines represent individual patients from time of admission until either their discharge (left) or death (right). Orange lines represent linear regression lines fitted on the linear scale. Vertical axis represents estimated daily probability of death during hospitalization (log-scale), which were obtained using the posterior median parameter values from the Markov model with *post-admission* regressors. Individual *a* was female aged over 80 with a history of hypertension and, generally, throughout the course of her stay was indicated to have a heightened risk of death: she was eventually discharged. Individual *b* was a male also aged over 80 with no recorded comorbidities: over time, his lab values changed substantially meaning their risk declined precipitously until his discharge. Individual *c* was female between 70 and 80 years old with a history of hypertension, hyperlipidemia, diabetes and coronary artery disease: over time, her values dramatically worsened, and she died five days after admission. Individual *d* was male aged between 50 and 60 years old and had end-stage renal disease, hypertension and diabetes: while his test values upon admission suggested low mortality risk, over time, his values worsened, and he died after more than 20 days in hospital. (B) Clinical decision tool for hospitalized COVID-19 patients: Each row of the diagram illustrates the key variables most useful for patient risk stratification based on the levels of available information. The block sizes, color and numbers within blocks represent odds ratios representing risk of daily mortality. The lab values indicate the cutoff point and direction of change associated with the given odds ratio.

### Dynamic triage

To facilitate decisions about risk stratification, we bring together the results of the Markov models into a clinical decision tool. This tool uses the most important factors for risk stratification dependent on the data available to clinicians at each time point. The decision tool is shown in Figure 4B.

## Discussion

In this retrospective cohort study, we calculated the daily mortality risk of patients hospitalized with COVID-19 by developing a Bayesian Markov model that uses patient characteristics, including demographic variables, comorbidities, biomarkers at admission and time-dependent biomarkers. Our results suggest that solely relying on admission variables had limited accuracy and that only a handful of factors contributed to predictive accuracy (in line with current evidence (29)). Contrastingly, incorporating into our model dynamic variation in biomarkers measured throughout hospitalization led to dramatic improvements in predictive power. The sequential approach we propose provides clinicians with a tool that enables decision making depending on the level of information available (30).

Since the beginning of the COVID-19 outbreak, numerous decision support tools for patient prognosis have been developed (16, 29). Given the reactive nature of the pandemic response, prognostic modelling efforts have suffered from a number of limitations, including lack of external validation and a high risk of bias (16). We fit our model to data from a single hospital in New York, where our study population mostly consists of black African American patients, which have been shown to be disparately affected by the COVID-19 pandemic (31, 32). Socio-economic variables can impact in-hospital mortality risk (9, 33) and external validation of ours and other models is therefore required. Finally, any clinical implementation requires an assessment of the impact of the prognostic model on clinicians’ behavior, patient health and associated costs (15).

Our model allows for risk stratification and triage. At the patient level, the model allows for individualization of care for each hospitalized COVID-19 patient and identification of need for additional levels of care (34); the main advantage of our approach is the incorporation of dynamic variables which allows for daily adjustments to the patient’s risk level. In addition, identification of high-risk patients and determining the surge capacity needed for advanced intensive care should allow for early resource allocation and ultimately improved outcomes for patients (6, 35). However, with insufficient surge capacity, triage should consider both patient prognosis and ethical considerations to avoid health inequities (36, 37).

A number of vaccines are in the process of undergoing regulatory approval or being rolled out. At this point, it is unclear what vaccination coverage will be achieved and the long-term immune protection conferred by vaccines is unknown (38). Currently, COVID-19 hospitalization rates remain high, and it is imperative that patients receive the best care available, and prognostic models like ours likely have a role to play in achieving this. Whilst our approach was applied to severely ill COVID-19 patients, similar patterns of inflammatory response and multi-organ injury are also seen in other acutely ill patients (39, 40). Identifying the effect of dynamic changes in relevant biomarkers on mortality risk in real time, using a tool like the one we develop, could be transformative in caring for these patients.

## Supporting information

Supplementary Methods and Results

## Data Availability

Per institutional guidelines, the data will not be shared publicly. Please contact the corresponding author if there is a need for data access.

