## Supplementary Methods and Results for "A Dynamic Bayesian Model for Identifying High-Mortality Risk in Hospitalized COVID-19 Patients"

### Characteristics of raw data:

| Test | Nobs | Mean | Median | 25% | 75% | Sd | Units |
| --- | --- | --- | --- | --- | --- | --- | --- |
| Albumin | 535 | 3.39 | 3.41 | 3.11 | 3.71 | 0.51 | g/dL |
| Alkaline Phosphatase | 535 | 81.58 | 69.00 | 56.83 | 89.25 | 57.73 | IU/L |
| AST | 535 | 80.46 | 42.00 | 28.00 | 69.50 | 257.97 | IU/L |
| BASO PCT | 464 | 0.50 | 0.40 | 0.30 | 0.60 | 0.39 | % |
| Bilirubin Total | 535 | 0.73 | 0.60 | 0.43 | 0.83 | 0.68 | mg/dL |
| Brain Natriuretic Peptide | 230 | 554.00 | 80.50 | 22.25 | 297.75 | 1491.03 | pg/mL |
| BUN | 539 | 39.19 | 26.50 | 16.00 | 50.17 | 36.21 | mg/dL |
| C Reactive Protein Serum | 421 | 163.14 | 149.00 | 80.00 | 246.00 | 104.04 | mg/L |
| Calcium Ionized Whole Blood | 466 | 1.12 | 1.11 | 1.06 | 1.17 | 0.10 | mmol/L |
| Calcium Level | 539 | 8.51 | 8.47 | 8.10 | 8.83 | 0.97 | mg/dL |
| CK | 154 | 724.08 | 212.50 | 112.75 | 612.00 | 1801.11 | ng/mL |
| CO2 | 539 | 22.45 | 23.00 | 20.00 | 25.00 | 4.58 | mmol/L |
| Creatine | 539 | 2.77 | 1.40 | 1.00 | 2.70 | 3.37 | mg/dL |
| EOS PCT | 464 | 0.62 | 0.30 | 0.10 | 0.80 | 0.88 | % |
| Ferritin | 403 | 2104.62 | 878.70 | 415.15 | 2132.30 | 4411.32 | ng/mL |
| Glucose Random | 539 | 187.04 | 144.00 | 112.25 | 229.50 | 116.67 | mg/dL |
| K Bld | 466 | 4.39 | 4.10 | 3.60 | 4.80 | 1.27 | mmol/L |
| Lactate Dehydrogenase | 411 | 564.78 | 468.00 | 342.00 | 637.50 | 431.16 | IU/L |
| LUC ABS | 464 | 0.16 | 0.10 | 0.10 | 0.20 | 0.11 | 10^3/mL |
| LUC PCT | 464 | 1.92 | 1.70 | 1.20 | 2.40 | 1.05 | % |
| LYM ABS | 464 | 1.00 | 0.90 | 0.60 | 1.20 | 0.58 | 10^3/µL |
| LYM PCT | 464 | 12.94 | 11.73 | 8.00 | 16.36 | 6.79 | % |
| MCH | 540 | 27.43 | 27.70 | 26.09 | 29.10 | 2.51 | pg |
| MCHC | 540 | 30.91 | 30.95 | 30.05 | 31.90 | 1.36 | % |
| MCV | 540 | 88.78 | 89.18 | 84.79 | 93.41 | 7.37 | fL |
| MONO ABS | 464 | 0.40 | 0.35 | 0.25 | 0.50 | 0.24 | 10^3/µL |
| MONO PCT | 464 | 4.98 | 4.40 | 3.30 | 6.15 | 2.44 | % |
| MPV | 540 | 9.81 | 9.60 | 8.90 | 10.45 | 1.24 | fL |
| Na Bld | 467 | 135.21 | 134.00 | 130.00 | 138.00 | 9.38 | mEq/L |
| NEU ABS | 482 | 7.30 | 6.20 | 4.33 | 8.94 | 4.38 | 10^3/µL |
| NEU PCT | 478 | 79.31 | 80.88 | 73.40 | 85.64 | 8.87 | % |
| PLAT | 540 | 232.06 | 205.00 | 150.00 | 299.25 | 111.92 | 10^3/µL |
| Procalcitonin (PCT) | 387 | 3.54 | 0.43 | 0.11 | 1.36 | 15.42 | ng/mL |
| Protein Total | 535 | 6.72 | 6.75 | 6.30 | 7.20 | 0.72 | g/dL |
| RBC | 540 | 4.49 | 4.52 | 4.00 | 4.99 | 0.79 | 10^6/µL |
| RDW | 540 | 14.87 | 14.43 | 13.60 | 15.61 | 1.70 | % |
| V pCO2 | 470 | 43.12 | 42.15 | 38.00 | 46.90 | 9.12 | KPa |
| WBC | 540 | 9.32 | 8.10 | 5.86 | 10.99 | 7.49 | 10^3/µL |

**Table S1: Summary of test values upon admission.** Here, “Nobs” indicates the number of patients who had measurements taken for each test on their first day of admission. Note, that the data here reflects summaries calculated after the steps outlined in the “Data cleaning” section and included n=540 individuals who had measurements taken on the first day. It includes data for those tests with 150 or more observations.

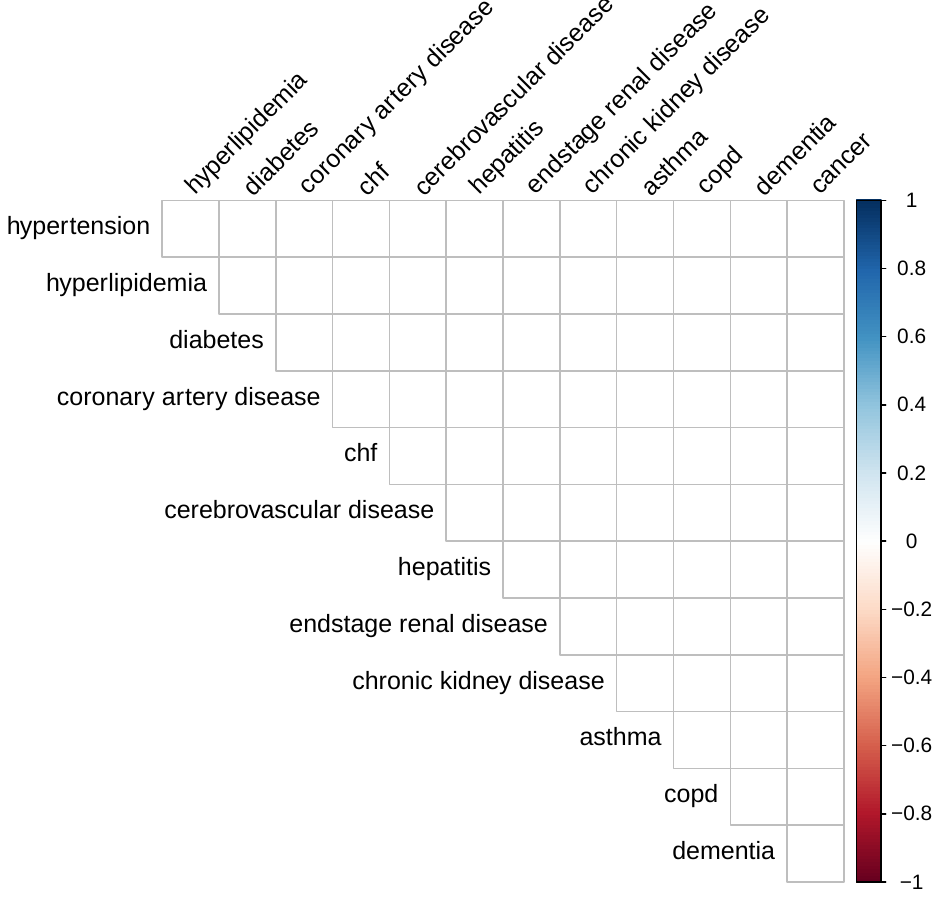

**Figure S1: Comorbidity correlations.** The color of each bubble indicates the sign and magnitude of the correlation in presence of paired correlations; the size of each bubble indicates the number of individuals with both conditions.

| Test | Discharged | Expired | Units |
| --- | --- | --- | --- |
| Albumin* | 3.46 (3.16-3.83) | 3.3 (3.04-3.61) | g/dL |
| Alkaline Phosphatase | 69 (57.5-88) | 70 (54-92) | IU/L |
| AST* | 37 (25-59.38) | 49 (35-84.5) | IU/L |
| BASO PCT* | 0.45 (0.3-0.6) | 0.4 (0.25-0.55) | % |
| Bilirubin Total* | 0.6 (0.4-0.8) | 0.67 (0.45-0.85) | mg/dL |
| Brain Natriuretic Peptide | 66 (14-351) | 88.5 (28.75-276) | pg/mL |
| BUN* | 23 (13.33-48) | 32.75 (20.62-56.38) | mg/dL |
| C Reactive Protein Serum* | 124 (61-208.5) | 192 (128-280.75) | mg/L |
| Calcium Ionized Whole Blood | 1.11 (1.06-1.17) | 1.11 (1.06-1.16) | mmol/L |
| Calcium Level | 8.5 (8.1-8.9) | 8.4 (8.05-8.8) | mg/dL |
| CK | 201 (88.25-560) | 242 (133.25-644.38) | ng/mL |
| CO2* | 23.5 (21-26) | 22 (19.5-24) | mmol/L |
| Creatine* | 1.25 (0.93-2.6) | 1.65 (1.15-2.84) | mg/dL |
| EOS PCT* | 0.4 (0.2-0.95) | 0.2 (0.1-0.46) | % |
| Ferritin* | 727.3 (371.3-1605.85) | 1173.75 (538.38-2708.27) | ng/mL |
| Glucose Random* | 133 (107-200) | 161.25 (121-262.88) | mg/dL |
| K Bld* | 4 (3.5-4.7) | 4.2 (3.8-4.91) | mmol/L |
| Lactate Dehydrogenase* | 427 (305.5-583) | 550.5 (416.62-757.25) | IU/L |
| LUC ABS* | 0.1 (0.1-0.2) | 0.1 (0.1-0.2) | 10^3/mL |
| LUC PCT* | 1.9 (1.3-2.6) | 1.45 (1-2) | % |
| LYM ABS* | 0.9 (0.7-1.25) | 0.8 (0.6-1.06) | 10^3/µL |
| LYM PCT* | 12.83 (8.74-17.39) | 10.43 (6.47-14.06) | % |
| MCH | 27.7 (26.1-29.04) | 27.62 (25.91-29.15) | pg |
| MCHC | 31.05 (30.15-31.9) | 30.88 (29.95-31.86) | % |
| MCV | 89.42 (85.15-92.99) | 89.1 (84.25-93.65) | fL |
| MONO ABS | 0.35 (0.25-0.5) | 0.35 (0.2-0.5) | 10^3/µL |
| MONO PCT* | 4.6 (3.5-6.5) | 3.92 (3-5.62) | % |
| MPV* | 9.42 (8.85-10.2) | 9.9 (9.3-10.66) | fL |
| Na Bld* | 133 (130-137) | 135 (130-141) | mEq/L |
| NEU ABS* | 5.8 (4-8.3) | 7.35 (4.9-10.1) | 10^3/µL |
| NEU PCT* | 79.18 (71.8-84.34) | 82.84 (78.81-88.4) | % |
| PLAT* | 222.75 (157.25-309.25) | 188 (143.75-260.38) | 10^3/µL |
| Procalcitonin (PCT)* | 0.28 (0.1-0.99) | 0.66 (0.24-2.06) | ng/mL |
| Protein Total | 6.8 (6.3-7.2) | 6.75 (6.26-7.2) | g/dL |
| RBC* | 4.49 (3.96-4.91) | 4.63 (4.07-5.1) | 10^6/µL |
| RDW | 14.32 (13.58-15.59) | 14.55 (13.8-15.84) | % |
| V pCO2* | 42.6 (38.85-47.45) | 40.6 (37.5-46) | KPa |
| WBC* | 7.76 (5.62-10.18) | 9.2 (6.7-12.24) | 10^3/µL |

**Table S2: Comparison of admission test values between discharged and expired patients.** The values represent the median test value in each group and, in parentheses, the interquartile ranges. Asterisks indicate p<0.05 in a Mann-Whitney U test. The table includes analyses for only those tests with 150 or more observations.

| Variable | Coef | OR | Std. Error (Coef) | z | p |
| --- | --- | --- | --- | --- | --- |
| Age * | 0.03189 | 1.032407 | 0.005673 | 5.622 | 1.89E-08 |
| Race (Black) | -6.73E-01 | 5.10E-01 | 1.01E+00 | -0.669 | 0.503 |
| Sex (Male) | 0.1301 | 1.139 | 0.1406 | 0.926 | 0.354 |
| Hypertension | 0.2724 | 1.3131 | 0.1574 | 1.73 | 0.0837 |
| Hyperlipidemia * | 0.3206 | 1.378 | 0.1626 | 1.972 | 0.0486 |
| Diabetes | -0.01543 | 0.98469 | 0.13999 | -0.11 | 0.912 |
| Coronary Artery Disease * | 0.655 | 1.9251 | 0.2183 | 3.001 | 0.00269 |
| CHF | -0.00112 | 0.998884 | 0.385469 | -0.003 | 0.998 |
| Cerebrovascular Disease | 0.301 | 1.3512 | 0.2683 | 1.122 | 0.262 |
| Hepatitis | 0.08392 | 1.08755 | 0.71201 | 0.118 | 0.906 |
| ESRD | -0.1576 | 0.8542 | 0.2684 | -0.587 | 0.557 |
| CKD | 0.4501 | 1.5685 | 0.3249 | 1.385 | 0.166 |
| Asthma | 0.03562 | 1.03626 | 0.36075 | 0.099 | 0.921 |
| COPD | -0.05242 | 0.94893 | 0.31025 | -0.169 | 0.866 |
| Dementia | 0.2922 | 1.3394 | 0.3612 | 0.809 | 0.419 |
| Cancer | 0.5188 | 1.6799 | 0.3253 | 1.595 | 0.111 |
| Albumin | -0.1427 | 0.867 | 0.1414 | -1.009 | 0.313 |
| Alkaline Phosphatase | 0.001818 | 1.001819 | 0.001063 | 1.71 | 0.0872 |
| AST | 0.000132 | 1.000132 | 0.000167 | 0.793 | 0.428 |
| BASO PCT | -0.1577 | 0.8541 | 0.2474 | -0.638 | 0.524 |
| Bilirubin Total * | 0.22284 | 1.24962 | 0.09665 | 2.306 | 0.0211 |
| Brain Natriuretic Peptide | 2.64E-05 | 1.00E+00 | 7.60E-05 | 0.348 | 0.728 |
| BUN * | 0.003683 | 1.00369 | 0.001529 | 2.409 | 0.016 |
| C Reactive Protein Serum * | 0.002519 | 1.002522 | 0.000709 | 3.551 | 0.000384 |
| Calcium Ionized Whole Blood | 0.1606 | 1.1742 | 0.7488 | 0.214 | 0.83 |
| Calcium Level | -0.03381 | 0.96675 | 0.09962 | -0.339 | 0.734 |
| CK | 4.591e-06 | 1.000e+00 | 4.976e-05 | 0.092 | 0.926 |
| CO2 * | -0.05639 | 0.94517 | 0.01458 | -3.868 | 0.00011 |
| Creatinine | 0.004869 | 1.00488 | 0.019792 | 0.246 | 0.806 |
| EOS PCT * | -0.3907 | 0.6766 | 0.1451 | -2.693 | 0.00708 |
| Ferritin * | 4.55E-05 | 1.00E+00 | 1.62E-05 | 2.807 | 0.005 |
| Glucose Random | 0.000208 | 1.000208 | 0.00061 | 0.341 | 0.733 |
| K Bld * | 0.10656 | 1.11245 | 0.04623 | 2.305 | 0.0212 |
| Lactate Dehydrogenase | 8.91E-05 | 1.00E+00 | 1.42E-04 | 0.626 | 0.532 |
| LUC ABS | -1.3806 | 0.2514 | 0.8219 | -1.68 | 0.093 |
| LUC PCT* | -0.2139 | 0.8074 | 0.09196 | -2.326 | 0.02 |
| LYM ABS | -0.3014 | 0.7397 | 0.1645 | -1.832 | 0.0669 |
| LYM PCT * | -0.0262 | 0.97414 | 0.01304 | -2.009 | 0.0446 |
| MCH | -0.02204 | 0.97821 | 0.03162 | -0.697 | 0.486 |
| MCHC* | -0.15524 | 0.85621 | 0.05647 | -2.749 | 0.00597 |
| MCV | 0.006799 | 1.006823 | 0.010629 | 0.64 | 0.522 |
| MONO ABS | -0.2446 | 0.783 | 0.3086 | -0.793 | 0.428 |
| MONO PCT | -0.0424 | 0.95849 | 0.03431 | -1.236 | 0.216 |
| MPV * | 0.14001 | 1.15028 | 0.05009 | 2.795 | 0.00519 |
| Na Bld * | 0.014981 | 1.015094 | 0.007163 | 2.091 | 0.0365 |
| NEU ABS | 0.02015 | 1.02036 | 0.01376 | 1.465 | 0.143 |
| NEU PCT * | 0.028528 | 1.028939 | 0.009965 | 2.863 | 0.0042 |
| PLAT * | -0.00254 | 0.997466 | 0.000716 | -3.546 | 0.000391 |
| Procalcitonin | 0.003304 | 1.003309 | 0.004132 | 0.8 | 0.424 |
| Protein Total | 0.11078 | 1.11715 | 0.09759 | 1.135 | 0.256 |
| RBC * | 0.26216 | 1.29974 | 0.09517 | 2.755 | 0.00588 |
| RDW * | 0.08475 | 1.08845 | 0.04177 | 2.029 | 0.0425 |
| V pCO2 | 0.010890 | 1.010950 | 0.008003 | 1.361 | 0.174 |
| WBC * | 0.012371 | 1.012447 | 0.006098 | 2.029 | 0.0425 |

**Table S3: Univariate Survival Analysis of baseline characteristics and admission test values.** The coefficients values are based on univariate Cox-regression analysis. The duration of survival is calculated from admission until either the patients were discharged or expired. Asterisks indicate p<0.05. The table includes analyses only for those tests with 150 or more observations.

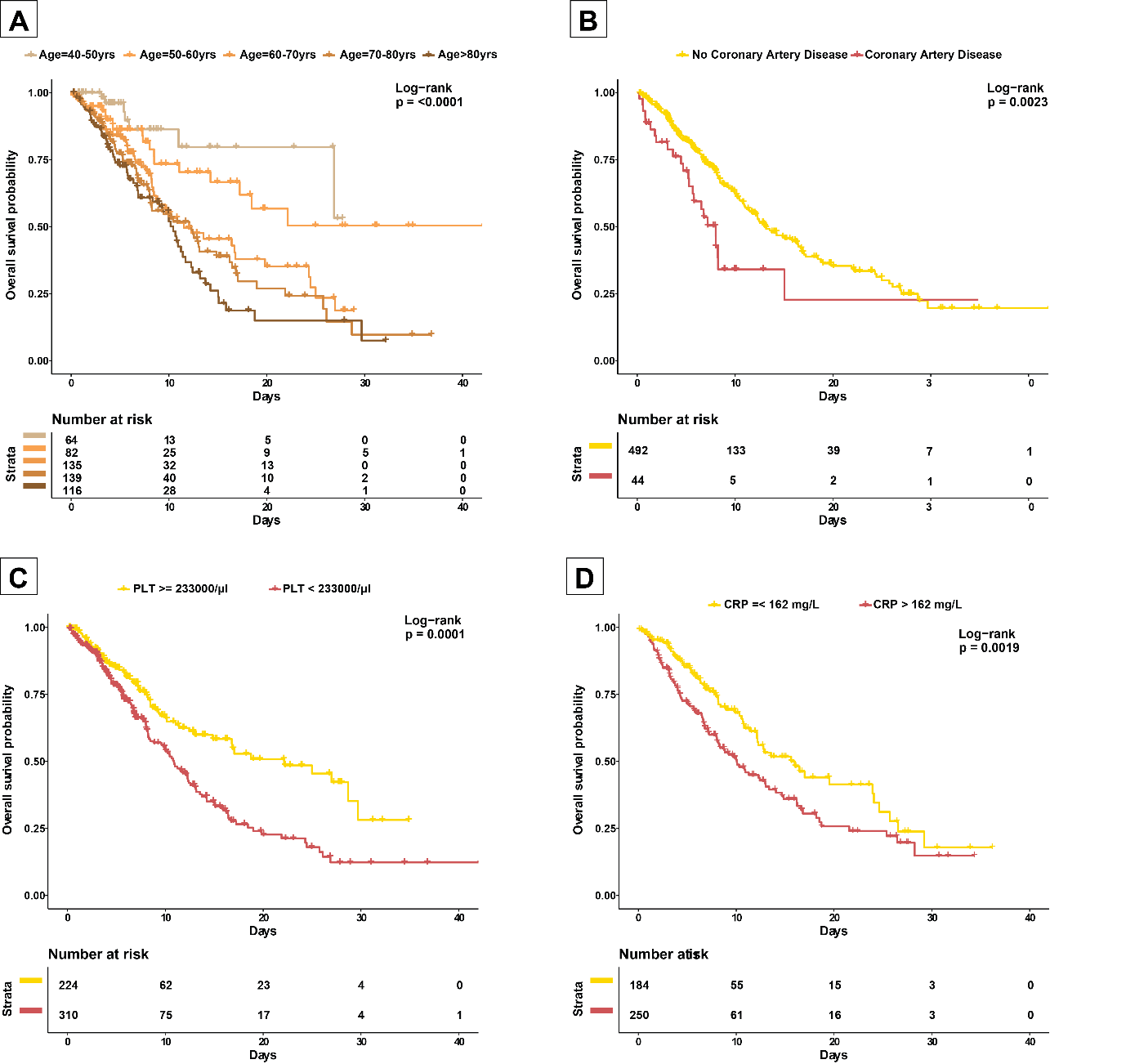

**Figure S2: Kaplan-Meier survival curves for select variables.** (A) Survival curves for various age-groups. (B) Survival curves for coronary artery disease status. (C) Survival curves for platelets level at admission. (D) Survival curves for CRP levels at admission.

| Variable | Univariate | Patient | Pat. + comorbidities | Admission | Post-admission |
| --- | --- | --- | --- | --- | --- |
| Age: 40 - 49 | 1.54 (0.73) | 0.79 (0.22) | 0.79 (0.20) | 0.71 (0.14) | 0.96 (0.40) |
| Age: 50 - 59 | 2.69 (0.94) | 0.98 (0.45) | 0.92 (0.33) | 0.93 (0.26) | 0.97 (0.40) |
| Age: 60 - 69 | 6.1 (1) | 1.94 (0.95) | 1.4 (0.87) | 1.02 (0.59) | 1.24 (0.78) |
| Age: 70 - 79 | 9.08 (1) | 2.94 (1) | 2.15 (0.98) | 1.08 (0.74) | 1.04 (0.6) |
| Age: 80+ | 9.78 (1) | 3.56 (1) | 2.78 (1) | 1.27 (0.86) | 0.99 (0.46) |
| Ethnicity: BAME | 1.92 (0.92) | 1.05 (0.62) | 1.04 (0.6) | 1 (0.47) | 1.02 (0.55) |
| Sex: male | 1.65 (1) | 1.47 (0.97) | 1.54 (0.98) | 1.05 (0.70) | 1.02 (0.57) |
| Admit: Saturday | 1.58 (0.89) | 1.1 (0.72) | 1.1 (0.72) | 1.02 (0.62) | 0.98 (0.44) |
| Admit: Sunday | 3.36 (1) | 2.14 (0.99) | 2.08 (0.99) | 2.08 (0.98) | 1.85 (0.9) |
| Admit: Monday | 1.61 (0.94) | 1.1 (0.74) | 1.13 (0.78) | 1.06 (0.7) | 1.03 (0.59) |
| Admit: Tuesday | 0.99 (0.49) | 0.89 (0.24) | 0.93 (0.28) | 0.97 (0.35) | 0.86 (0.29) |
| Admit: Wednesday | 1.16 (0.67) | 0.94 (0.32) | 0.93 (0.30) | 0.93 (0.27) | 1.01 (0.52) |
| Admit: Thursday | 1.16 (0.68) | 1 (0.52) | 1 (0.48) | 1 (0.54) | 1.08 (0.66) |
| Days since first COVID patient admitted to hospital | 0.76 (0.00) | 0.85 (0.06) | 0.83 (0.04) | 0.85 (0.08) | 0.85 (0.19) |
| Hypertension | 1.88 (1.00) | . | 1.67 (0.98) | 1.55 (0.95) | 1.26 (0.79) |
| Hyperlipidemia | 1.76 (0.99) | . | 1.11 (0.76) | 1.12 (0.79) | 1.1 (0.68) |
| Diabetes | 1.55 (0.99) | . | 1.11 (0.78) | 1.12 (0.80) | 1.07 (0.66) |
| Coronary Artery Disease | 2.18 (0.99) | . | 1.2 (0.8) | 1.14 (0.79) | 1.98 (0.89) |
| CHF | 0.68 (0.21) | . | 0.91 (0.29) | 0.97 (0.38) | 0.9 (0.33) |
| Cerebrovascular Disease | 2.78 (0.99) | . | 1.6 (0.91) | 1.05 (0.68) | 1.47 (0.81) |
| Hepatitis | 1.48 (0.67) | . | 1.02 (0.56) | 1 (0.51) | 1 (0.49) |
| Endstage Renal Disease | 0.61 (0.06) | . | 0.94 (0.33) | 1 (0.46) | 1 (0.5) |
| Chronic Kidney Disease | 1.59 (0.84) | . | 0.98 (0.44) | 0.98 (0.39) | 0.79 (0.26) |
| Asthma | 0.84 (0.34) | . | 1 (0.52) | 1 (0.53) | 0.99 (0.48) |
| COPD | 1.26 (0.72) | . | 1.1 (0.71) | 1.02 (0.62) | 1.17 (0.71) |
| Dementia | 2.54 (0.95) | . | 1.41 (0.83) | 1.01 (0.58) | 1.02 (0.54) |
| Cancer | 3.21 (0.99) | . | 1.26 (0.80) | 1.09 (0.73) | 1.57 (0.82) |
| Albumin | 0.72 (0.00) | . | . | 0.68 (0.02) | 0.78 (0.16) |
| Alkaline phosphatase | 1.1 (0.84) | . | . | 1.05 (0.77) | 1.21 (0.9) |
| AST | 1.38 (1) | . | . | 1.05 (0.77) | 1.06 (0.66) |
| BASO PCT | 0.94 (0.26) | . | . | 1 (0.52) | 0.99 (0.45) |
| Bilirubin Total | 1.31 (0.99) | . | . | 1.13 (0.88) | 1.48 (0.94) |
| BUN | 1.34 (1) | . | . | 1.08 (0.79) | 1.74 (0.98) |
| Calcium Level | 0.72 (0) | . | . | 0.92 (0.21) | 0.97 (0.38) |
| CO2 | 0.75 (0) | . | . | 0.97 (0.32) | 0.92 (0.27) |
| Creatine | 0.94 (0.27) | . | . | 0.93 (0.21) | 0.93 (0.31) |
| EOS PCT | 0.47 (0) | . | . | 0.74 (0.04) | 0.76 (0.1) |
| Glucose Random | 1.26 (0.99) | . | . | 1.03 (0.68) | 1.42 (0.94) |
| LUC ABS | 0.91 (0.18) | . | . | 0.99 (0.43) | 0.94 (0.33) |
| LUC PCT | 0.65 (0) | . | . | 0.97 (0.34) | 0.89 (0.28) |
| LYM ABS | 0.7 (0) | . | . | 0.95 (0.28) | 0.98 (0.42) |
| LYM PCT | 0.56 (0) | . | . | 0.97 (0.36) | 1.01 (0.53) |
| MCH | 0.95 (0.27) | . | . | 1 (0.49) | 1.04 (0.61) |
| MCHC | 0.9 (0.14) | . | . | 0.98 (0.36) | 0.98 (0.43) |
| MCV | 0.98 (0.43) | . | . | 1 (0.51) | 1.08 (0.68) |
| MONO ABS | 1.06 (0.73) | . | . | 1 (0.5) | 0.89 (0.28) |
| MONO PCT | 0.73 (0) | . | . | 1 (0.55) | 0.88 (0.28) |
| MPV | 1.42 (1) | . | . | 1.05 (0.75) | 1.17 (0.79) |
| NEU ABS | 1.6 (1) | . | . | 1.1 (0.78) | 1.24 (0.75) |
| NEU PCT | 2 (1) | . | . | 1.33 (0.89) | 2.23 (0.94) |
| PLAT | 0.74 (0) | . | . | 0.7 (0.01) | 0.66 (0.06) |
| Protein Total | 1.06 (0.71) | . | . | 1.43 (0.98) | 1.69 (0.96) |
| RBC | 1.32 (1) | . | . | 1.3 (0.97) | 1.32 (0.89) |
| RDW | 1.1 (0.86) | . | . | 1.12 (0.85) | 1.52 (0.95) |
| WBC | 1.41 (1) | . | . | 1.01 (0.55) | 1.4 (0.82) |
| Albumin %Δ | 0.47 (0) | . | . | . | 0.82 (0.19) |
| Alkaline Phosphatase %Δ | 4.88 (1) | . | . | . | 1.24 (0.82) |
| AST %Δ | 253.5 (1) | . | . | . | 1.28 (0.79) |
| BASO PCT %Δ | 0.52 (0) | . | . | . | 0.6 (0.06) |
| Bilirubin Total %Δ | 3.7 (1) | . | . | . | 2.63 (0.96) |
| BUN %Δ | 11.07 (1) | . | . | . | 8.66 (1) |
| Calcium Level %Δ | 0.74 (0) | . | . | . | 0.91 (0.29) |
| CO2 %Δ | 0.33 (0) | . | . | . | 0.59 (0.04) |
| Creatine %Δ | 22.41 (1) | . | . | . | 1.08 (0.64) |
| EOS PCT %Δ | 0.41 (0) | . | . | . | 0.87 (0.23) |
| Glucose Random %Δ | 1.6 (1) | . | . | . | 1.32 (0.92) |
| LUC ABS %Δ | 1.53 (1) | . | . | . | 1.4 (0.87) |
| LUC PCT %Δ | 0.55 (0) | . | . | . | 0.82 (0.22) |
| LYM ABS %Δ | 0.77 (0.01) | . | . | . | 0.99 (0.46) |
| LYM PCT %Δ | 0.18 (0) | . | . | . | 0.27 (0.01) |
| MCH %Δ | 0.89 (0.12) | . | . | . | 1.01 (0.52) |
| MCHC %Δ | 0.5 (0) | . | . | . | 0.73 (0.16) |
| MCV %Δ | 2.33 (1) | . | . | . | 1.55 (0.9) |
| MONO ABS %Δ | 1.16 (0.94) | . | . | . | 0.88 (0.26) |
| MONO PCT %Δ | 0.38 (0) | . | . | . | 0.85 (0.23) |
| MPV %Δ | 2.31 (1) | . | . | . | 1.44 (0.9) |
| NEU ABS %Δ | 3.73 (1) | . | . | . | 1.22 (0.76) |
| NEU PCT %Δ | 3.8 (1) | . | . | . | 1.24 (0.79) |
| PLAT %Δ | 0.66 (0) | . | . | . | 0.59 (0.04) |
| Protein Total %Δ | 0.63 (0) | . | . | . | 1.16 (0.76) |
| RBC %Δ | 0.55 (0) | . | . | . | 0.7 (0.09) |
| RDW %Δ | 1.8 (1) | . | . | . | 1.03 (0.61) |
| WBC %Δ | 4 (1) | . | . | . | 1.64 (0.91) |
| Number of patients | Varies | 516 | 516 | 475 | 475 |

**Table S4: Logistic model results.** Dependent variable is binary indicator representing whether or not a patient died. Unparenthesised values indicate posterior median odds ratios; values in parentheses indicate the posterior probability that the odds ratio exceeds zero. For a description of the methods, see the “Determining patient risk” section.

| Variable | Patient | Pat. + comorbidities | Admission | Post-admission |
| --- | --- | --- | --- | --- |
| Age: 40 - 49 | 0.77 (0.25) | 0.75 (0.23) | 0.57 (0.1) | 0.69 (0.20) |
| Age: 50 - 59 | 0.69 (0.16) | 0.62 (0.11) | 0.59 (0.08) | 0.22* (0.0) |
| Age: 60 - 69 | 1.48 (0.88) | 1.30 (0.79) | 1.06 (0.57) | 0.98 (0.48) |
| Age: 70 - 79 | 1.64 (0.94) | 1.43 (0.86) | 1.18 (0.69) | 0.9 (0.37) |
| Age: 80+ | 2.07* (0.99) | 1.88* (0.96) | 1.64 (0.92) | 1.15 (0.66) |
| Sex: male | 1.23 (0.91) | 1.33* (0.96) | 1.04 (0.57) | 1.00 (0.51) |
| Ethnicity: BAME | 0.56 (0.07) | 0.54 (0.06) | 0.38* (0.02) | 0.49 (0.06) |
| Days since first COVID patient admitted to hospital | 0.90 (0.10) | 0.91 (0.11) | 0.85 (0.06) | 0.84 (0.09) |
| Admit: Sunday | 1.31 (0.80) | 1.28 (0.77) | 1.27 (0.76) | 1.22 (0.72) |
| Admit: Monday | 0.92 (0.40) | 0.94 (0.42) | 1.02 (0.52) | 0.87 (0.34) |
| Admit: Tuesday | 0.44* (0.01) | 0.44* (0.01) | 0.42* (0.01) | 0.23* (0.00) |
| Admit: Wednesday | 0.58 (0.05) | 0.57 (0.05) | 0.49* (0.03) | 0.41* (0.02) |
| Admit: Thursday | 0.91 (0.38) | 0.9 (0.36) | 0.86 (0.33) | 0.97 (0.46) |
| Admit: Friday | 0.83 (0.27) | 0.82 (0.26) | 0.75 (0.19) | 0.72 (0.18) |
| Admit: Saturday | 0.85 (0.32) | 0.89 (0.36) | 0.95 (0.44) | 0.81 (0.3) |
| Hypertension | . | 1.37 (0.95) | 1.34 (0.92) | 1.15 (0.73) |
| Hyperlipidemia | . | 1.1 (0.71) | 1.27 (0.87) | 1.15 (0.71) |
| Diabetes | . | 0.92 (0.29) | 0.97 (0.43) | 0.83 (0.19) |
| Coronary Artery Disease (CAD) | . | 1.57* (0.95) | 1.77* (0.98) | 2.62* (1.00) |
| CHF | . | 0.91 (0.39) | 0.67 (0.17) | 0.67 (0.19) |
| Cerebrovascular Disease | . | 1.27 (0.76) | 1.35 (0.80) | 1.77 (0.91) |
| Hepatitis | . | 0.88 (0.41) | 0.92 (0.43) | 1.08 (0.56) |
| End-stage Renal Disease | . | 1.01 (0.51) | 1.18 (0.69) | 1.04 (0.54) |
| Chronic Kidney Disease | . | 0.93 (0.42) | 0.9 (0.4) | 0.94 (0.44) |
| Asthma | . | 1.07 (0.58) | 1.04 (0.54) | 1.22 (0.69) |
| COPD | . | 1.18 (0.71) | 1.33 (0.81) | 2.19* (0.97) |
| Dementia | . | 0.95 (0.44) | 0.89 (0.39) | 0.82 (0.32) |
| Cancer | . | 1.09 (0.61) | 0.92 (0.41) | 0.91 (0.39) |
| Albumin %Δ | . | . | . | 0.85 (0.15) |
| Albumin | . | . | 0.79* (0.04) | 0.79 (0.07) |
| Alkaline Phosphatase %Δ | . | . | . | 1.17* (0.97) |
| Alkaline Phosphatase | . | . | 1.09 (0.84) | 1.15 (0.91) |
| AST %Δ | . | . | . | 1.06 (0.89) |
| AST | . | . | 1.04 (0.72) | 1.04 (0.68) |
| Baso PCT %Δ | . | . | . | 0.98 (0.44) |
| Baso PCT | . | . | 1.04 (0.63) | 0.93 (0.31) |
| Bilirubin Total %Δ | . | . | . | 1.11 (0.95) |
| Bilirubin Total | . | . | 1.17 (0.92) | 1.20 (0.92) |
| BUN %Δ | . | . | . | 1.15 (0.84) |
| BUN %Δ | . | . | 1.07 (0.69) | 1.11 (0.77) |
| Calcium Level %Δ | . | . | . | 1.10 (0.80) |
| Calcium Level | . | . | 0.87 (0.18) | 1.03 (0.57) |
| CO2 %Δ | . | . | . | 0.73* (0.01) |
| CO2 | . | . | 0.93 (0.25) | 0.88 (0.17) |
| Creatine %Δ | . | . | . | 1.11 (0.82) |
| Creatine | . | . | 0.90 (0.23) | 1.01 (0.52) |
| EOS PCT %Δ | . | . | . | 0.62* (0.00) |
| EOS PCT | . | . | 0.76* (0.03) | 0.85 (0.15) |
| Glucose Random %Δ | . | . | . | 1.00 (0.51) |
| Glucose Random | . | . | 1.06 (0.73) | 1.18 (0.91) |
| LUC Abs %Δ | . | . | . | 1.25* (0.95) |
| LUC Abs | . | . | 1.05 (0.62) | 1.18 (0.78) |
| LUC PCT %Δ | . | . | . | 0.90 (0.28) |
| LUC PCT | . | . | 0.99 (0.48) | 0.92 (0.36) |
| LYM Abs %Δ | . | . | . | 1.14 (0.8) |
| LYM Abs | . | . | 1.02 (0.54) | 1.09 (0.65) |
| LYM PCT %Δ | . | . | . | 0.69 (0.07) |
| LYM PCT | . | . | 1.17 (0.72) | 1.72 (0.94) |
| MCH %Δ | . | . | . | 1.06 (0.58) |
| MCH | . | . | 1.32 (0.72) | 1.15 (0.62) |
| MCHC %Δ | . | . | . | 0.73 (0.2) |
| MCHC | . | . | 0.78 (0.15) | 0.69 (0.1) |
| MCV %Δ | . | . | . | 1.01 (0.52) |
| MCV | . | . | 0.89 (0.4) | 1.14 (0.61) |
| MONO Abs %Δ | . | . | . | 0.91 (0.32) |
| MONO Abs | . | . | 0.80 (0.13) | 0.75 (0.12) |
| MONO PCT %Δ | . | . | . | 0.92 (0.34) |
| MONO PCT | . | . | 1.26 (0.89) | 1.32 (0.88) |
| MPV %Δ | . | . | . | 1.16 (0.92) |
| MPV | . | . | 0.92 (0.2) | 0.9 (0.19) |
| NEU Abs %Δ | . | . | . | 0.84 (0.20) |
| NEU Abs | . | . | 1.16 (0.7) | 0.8 (0.26) |
| NEU PCT %Δ | . | . | . | 1.06 (0.61) |
| NEU PCT | . | . | 1.55 (0.92) | 2.48* (0.99) |
| PLAT (PLTs) %Δ | . | . | . | 0.69* (0.01) |
| PLAT (PLTs) | . | . | 0.78* (0.03) | 0.85 (0.14) |
| Protein Total %Δ | . | . | . | 1.25 (0.92) |
| Protein Total | . | . | 1.32* (0.99) | 1.18 (0.88) |
| RBC %Δ | . | . | . | 0.94 (0.33) |
| RBC | . | . | 1.41* (1.00) | 1.68* (1.00) |
| RDW %Δ | . | . | . | 1.28* (0.99) |
| RDW | . | . | 1.25* (0.96) | 1.46* (0.99) |
| WBC %Δ | . | . | . | 1.5* (0.99) |
| WBC | . | . | 0.89 (0.31) | 1.3 (0.83) |
| Time since patient admitted | . | . | . | 1.36* (0.97) |
| Number of patients | 475 | 475 | 475 | 475 |

**Table S5: Markov model results.** Unparenthesised values indicate posterior median odds ratios associated with daily mortality risk; values in parentheses indicate the posterior probability that the odds ratio exceeds zero. For a description of the methods, see the “Determining patient risk” section.

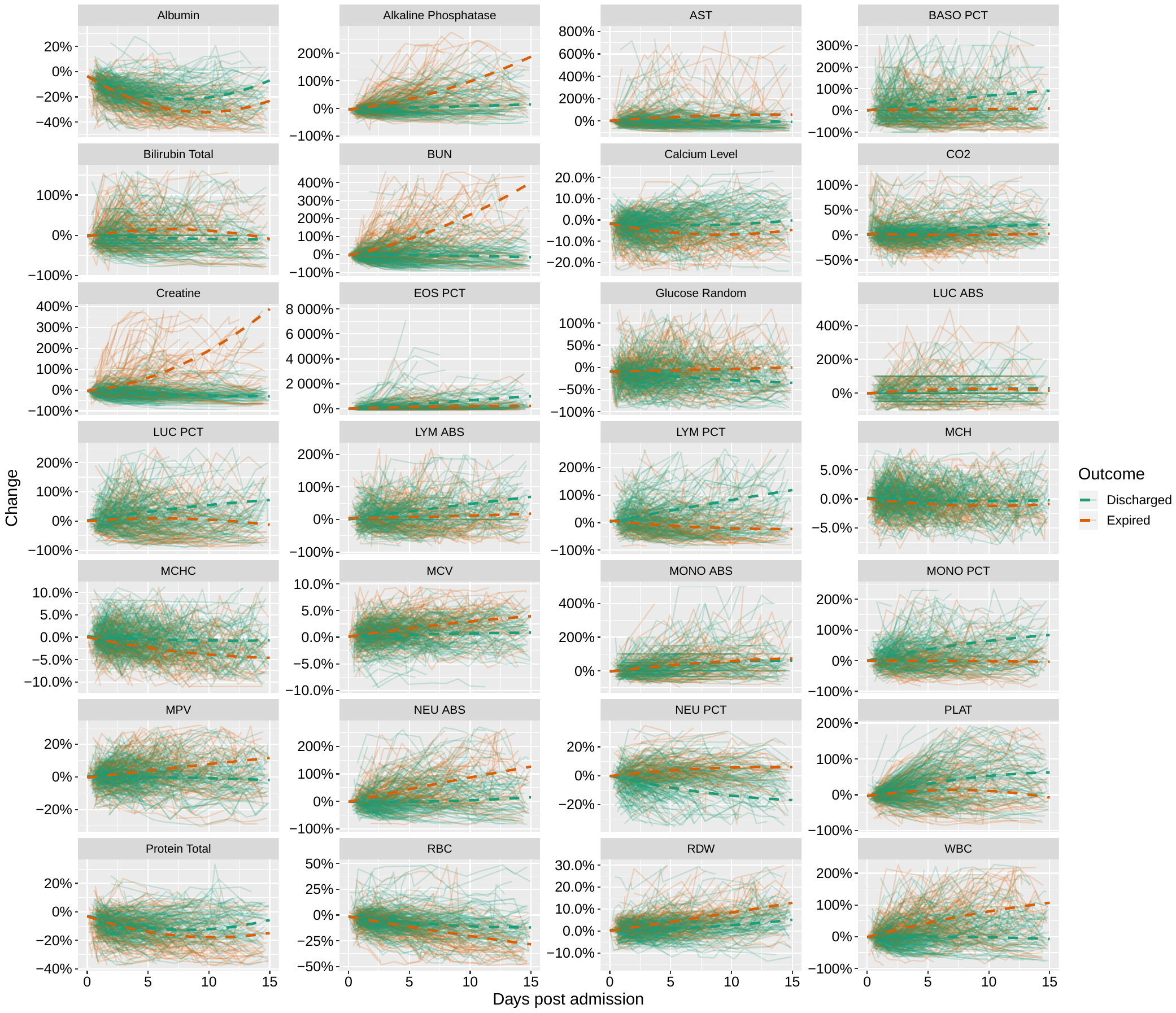

**Figure S3: Trends in lab values.** Solid lines indicate individual patient trajectories; dashed lines indicate fitted regression lines (see “Trend analysis” section for further information). Vertical axes show the percentage change in test values from their values upon admission.
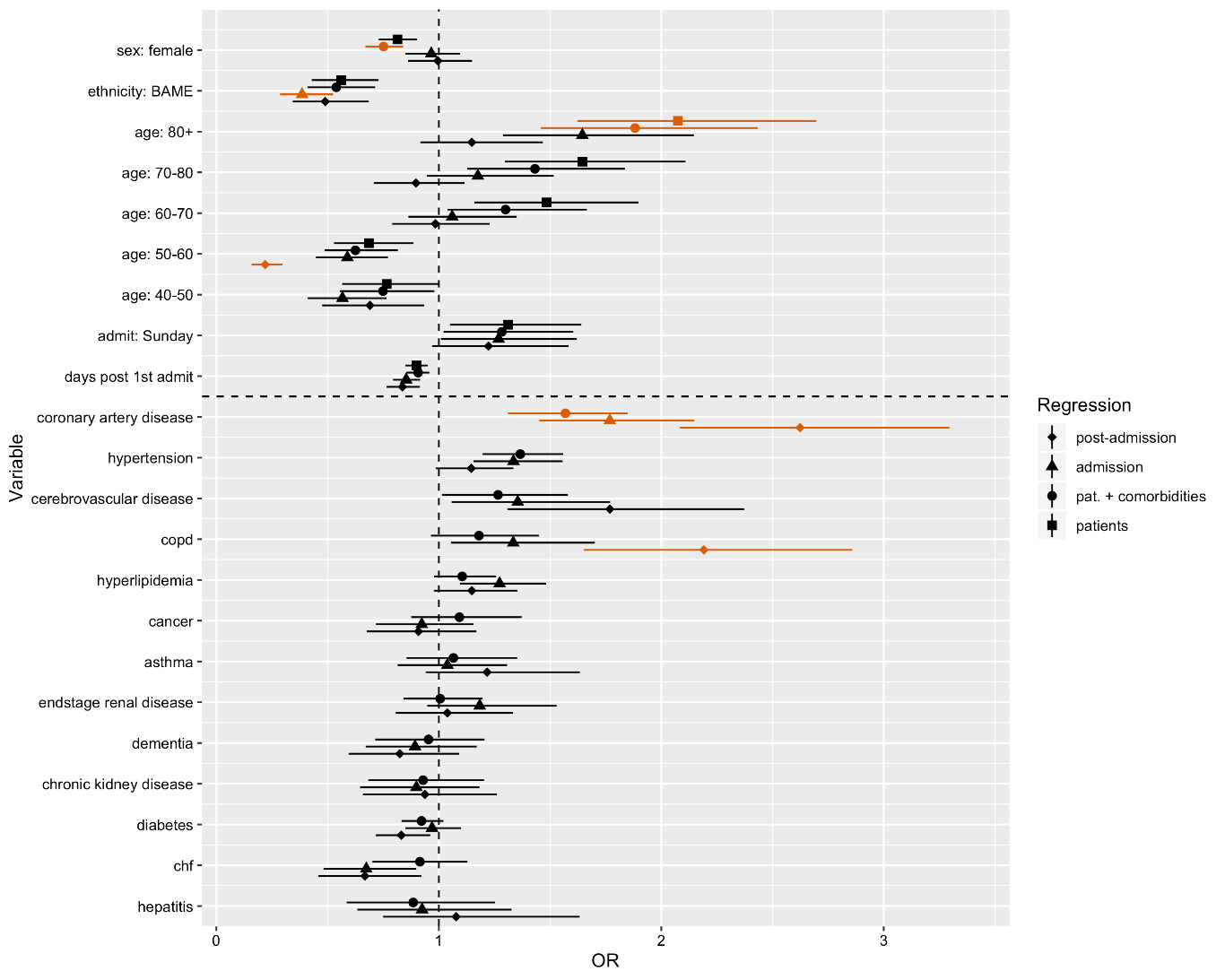

**Figure S4: Markov model: estimated odds ratios indicating mortality risk.** Each marker represents a different selection of variables included as regressors (see “Determining patient risk” section). The upper and lower whiskers indicate the 75% and 25% posterior quantiles; the middle points of each range indicate the posterior median. Those points shown in orange indicate that the 5%-95% posterior quantiles did not cross zero.

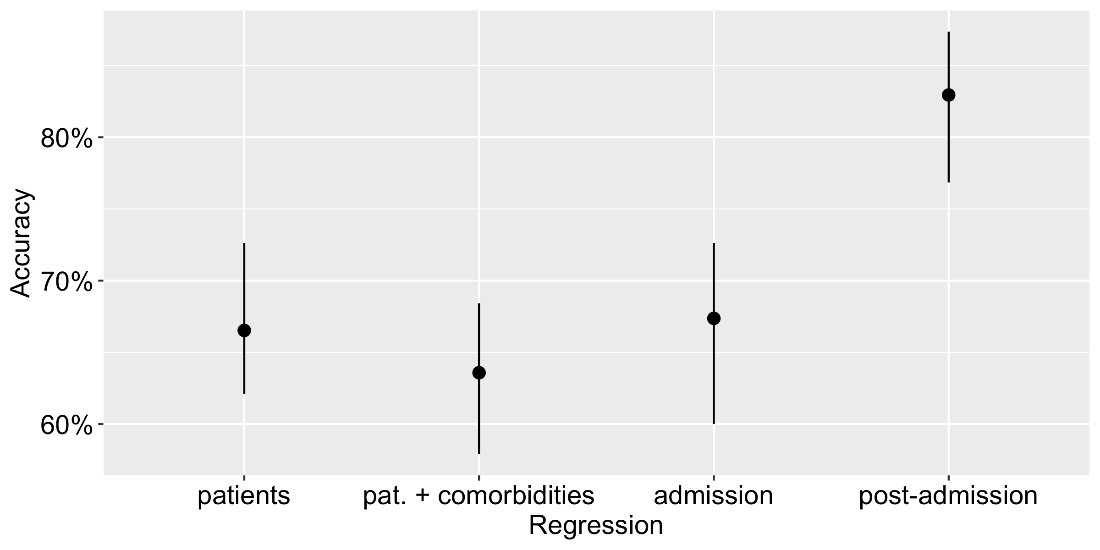

**Figure S5: Markov model: predictive accuracy.** The accuracy of each regression was assessed using cross-validation as described in the “Markov model checking using cross-validation" section. The upper and lower whiskers indicate the minimum and maximum predictive accuracy achieved across all independent testing sets. The middle point indicates the mean accuracy.

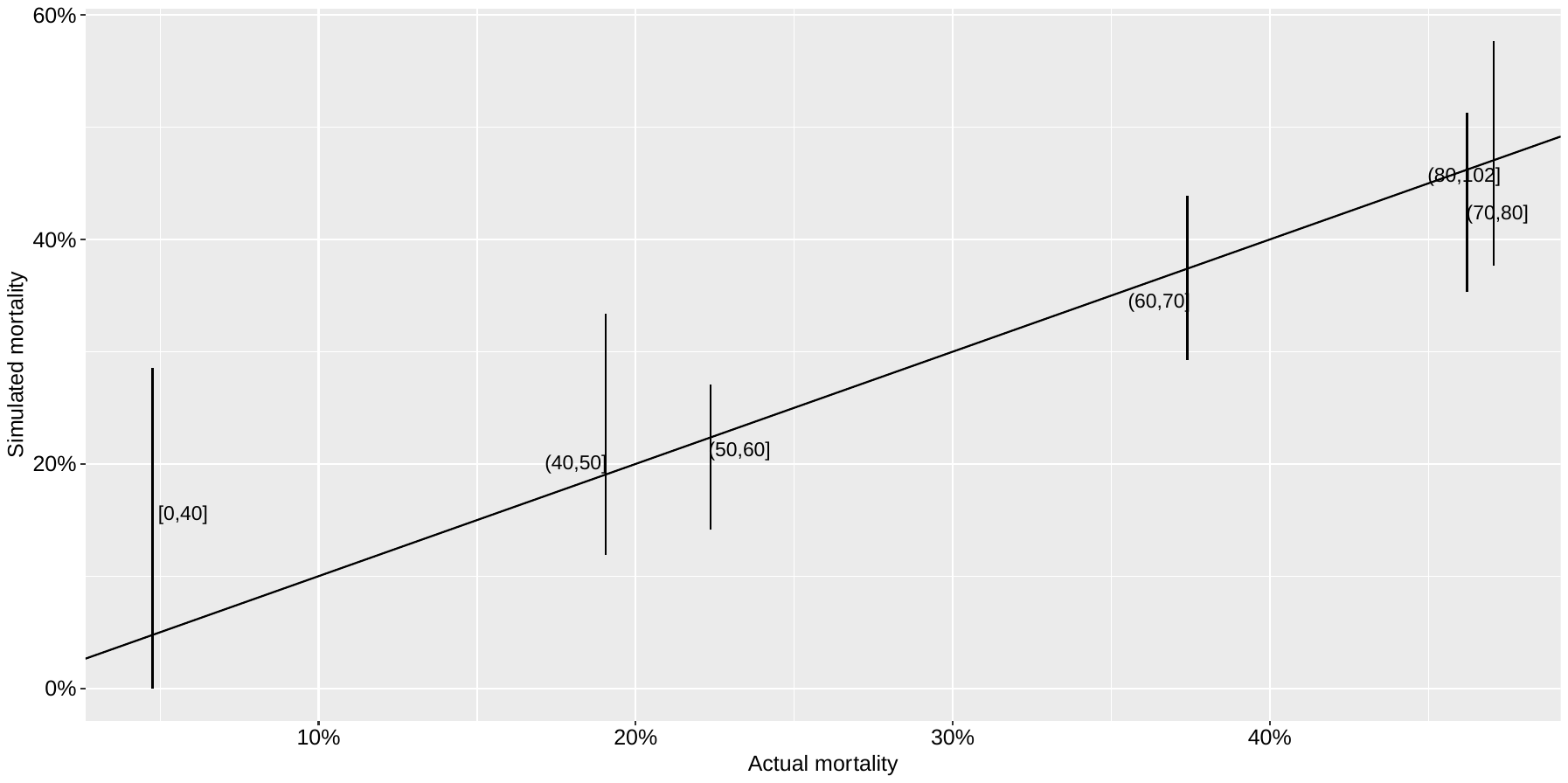

**Figure S6: Posterior predictive check: simulated versus actual mortality by age group.** The labels indicate each age group. Upper and lower whiskers show 97.5% and 2.5% posterior quantiles; points indicate posterior medians. These plots were produced using the *post-admission* Markov model.

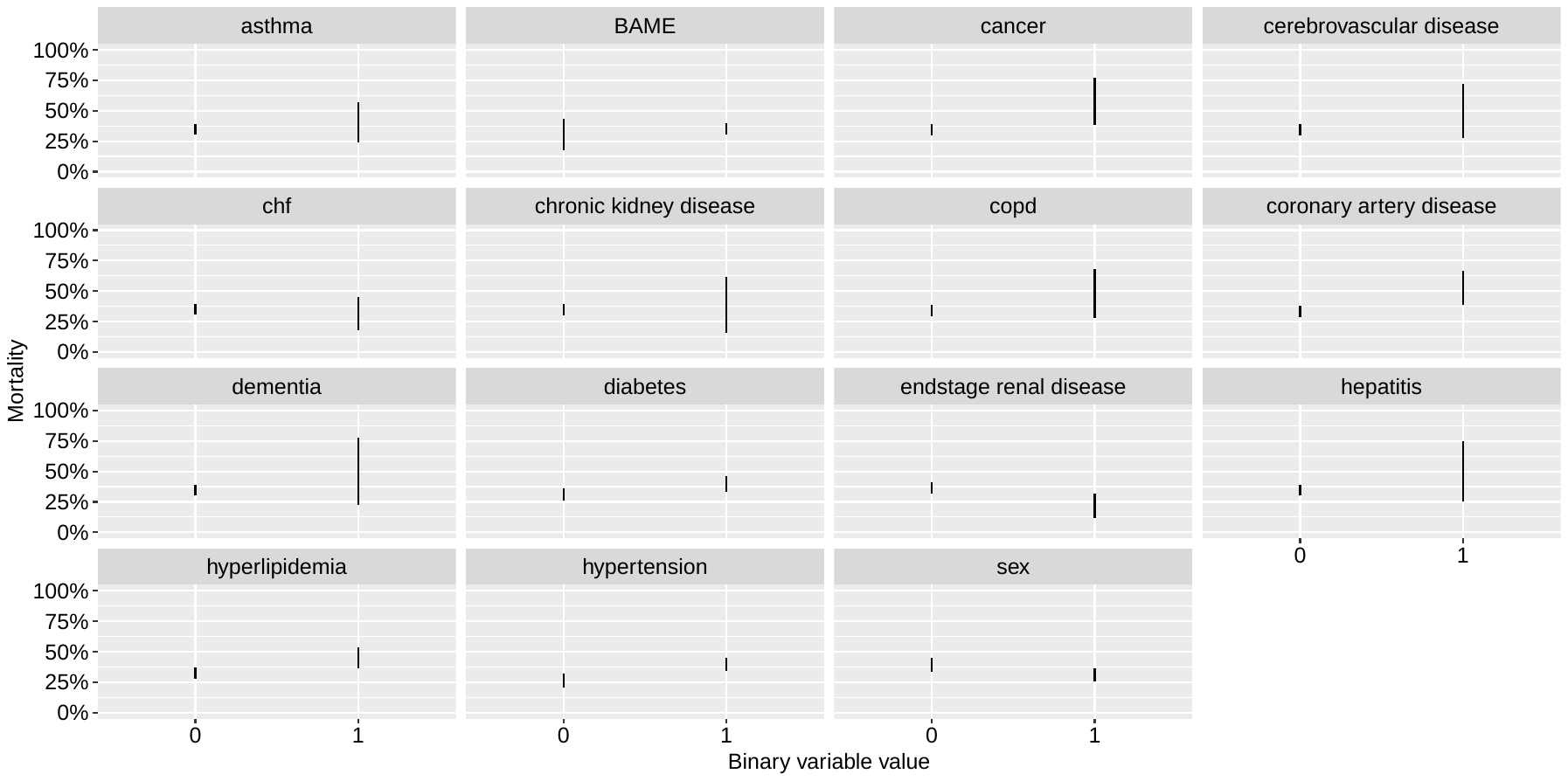

**Figure S7:** **Posterior predictive check: simulated (black) and actual (orange) mortality across different subgroups.** Upper and lower whiskers show 97.5% and 2.5% posterior quantiles; points indicate posterior medians. These plots were produced using the *post-admission* Markov model.

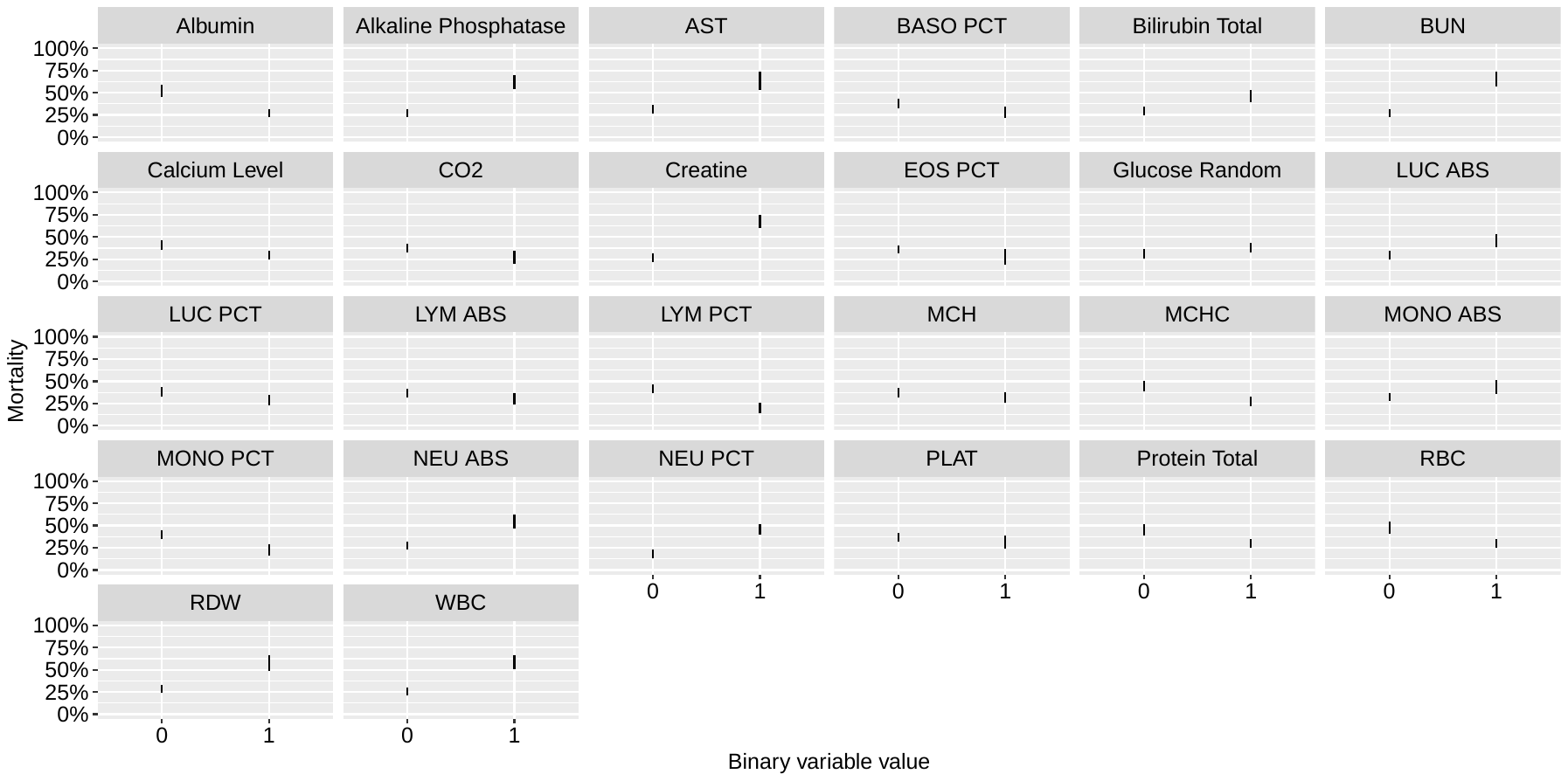

**Figure S8:** **Posterior predictive check: simulated (black) and actual (orange) mortality according to last observed change in lab values.** The binary indicator shows whether or not the mean observed change in lab values for a patient was above or below zero (on a standardised scale). Upper and lower whiskers show 97.5% and 2.5% posterior quantiles; points indicate posterior medians. These plots were produced using the *post-admission* Markov model.

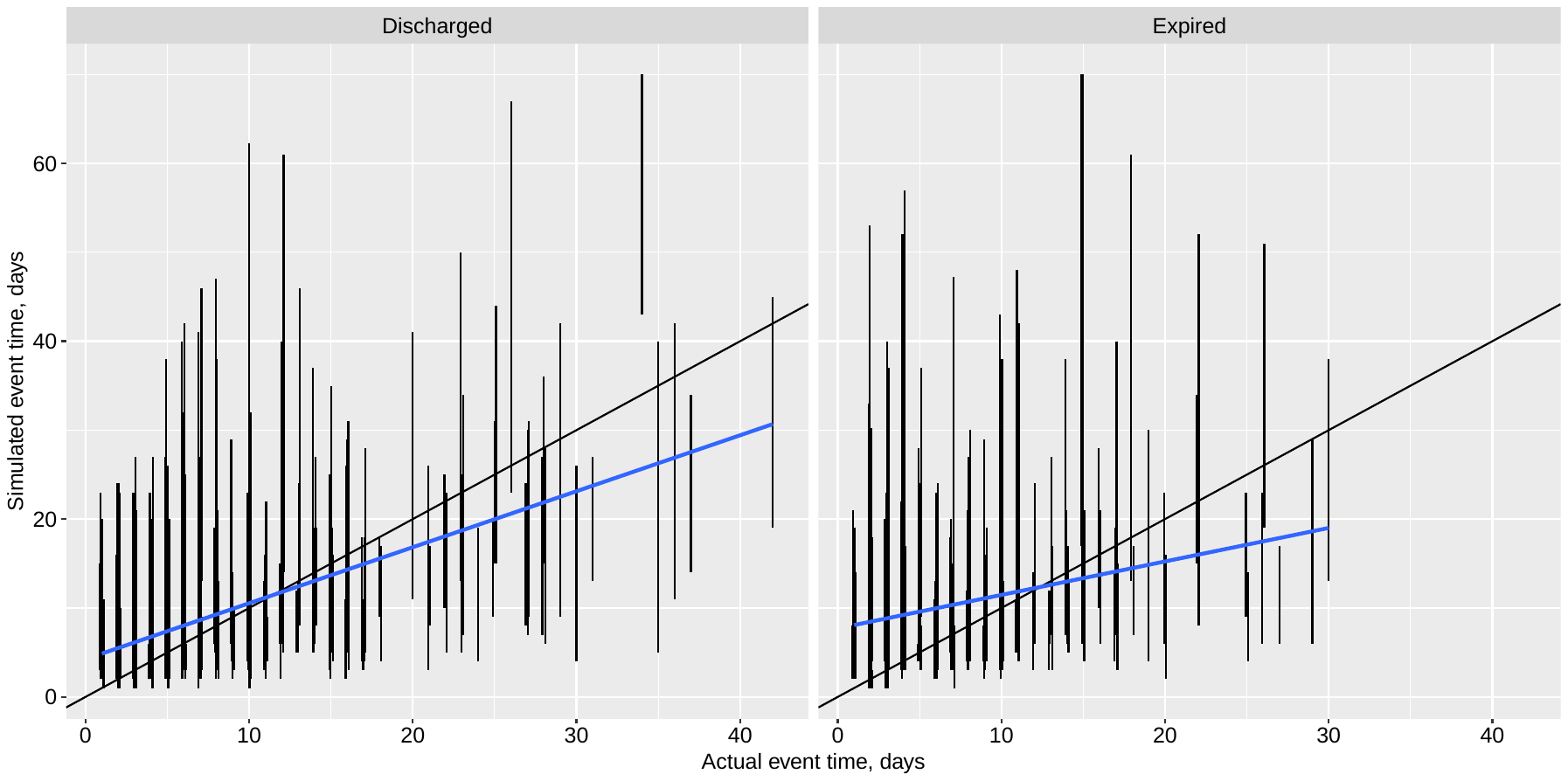

**Figure S9:** **Posterior predictive check: simulated versus actual hospitalization duration.** Upper and lower whiskers show 97.5% and 2.5% posterior quantiles; points indicate posterior medians. These plots were produced using the *post-admission* Markov model. The black diagonal lines show the actual=simulated values; the blue lines show the linear regression fit.

|  | Initial values | | | Percentage changes | |
| --- | --- | --- | --- | --- | --- |
| Test | **Mean** | **SD** | **Unit** | **Mean** | **SD** |
| Albumin | 3.40 | 0.49 | g/dL | -13.80% | 13.90% |
| Alkaline Phosphatase | 82.18 | 58.95 | IU/L | 23.26% | 76.40% |
| AST | 79.51 | 261.75 | IU/L | 61.69% | 714.32% |
| BASO PCT | 0.51 | 0.40 | % | 23.24% | 126.28% |
| Bilirubin Total | 0.74 | 0.70 | mg/dL | 5.21% | 59.71% |
| BUN | 37.68 | 32.96 | mg/dL | 31.14% | 126.88% |
| Calcium Level | 8.50 | 0.98 | mg/dL | -1.20% | 8.10% |
| CO2 | 22.61 | 4.40 | mmol/L | 5.99% | 24.86% |
| Creatine | 2.71 | 3.26 | mg/dL | 12.75% | 105.02% |
| EOS PCT | 0.64 | 0.94 | % | 337.86% | 835.22% |
| Glucose Random | 188.28 | 117.95 | mg/dL | -7.99% | 51.62% |
| LUC ABS | 0.16 | 0.11 | 10^3/mL | 23.79% | 80.67% |
| LUC PCT | 1.94 | 1.09 | % | 16.89% | 73.38% |
| LYM ABS | 0.99 | 0.58 | 10^3/µL | 18.82% | 61.29% |
| LYM PCT | 12.75 | 7.02 | % | 15.76% | 71.94% |
| MCH | 27.43 | 2.45 | pg | 0.04% | 3.43% |
| MCHC | 30.93 | 1.37 | % | -0.84% | 4.15% |
| MCV | 88.76 | 7.18 | fL | 1.01% | 2.76% |
| MONO ABS | 0.41 | 0.25 | 10^3/µL | 36.93% | 99.95% |
| MONO PCT | 4.97 | 2.45 | % | 19.68% | 57.48% |
| MPV | 9.77 | 1.21 | fL | 1.21% | 10.15% |
| NEU ABS | 7.41 | 4.54 | 10^3/µL | 20.11% | 67.51% |
| NEU PCT | 79.37 | 9.00 | % | -1.82% | 11.19% |
| PLAT (PLTs) | 236.15 | 112.60 | 10^3/µL | 25.05% | 61.24% |
| Protein Total | 6.73 | 0.68 | 10^3/µL | -7.19% | 10.58% |
| RBC | 4.47 | 0.78 | g/dL | -7.80% | 14.41% |
| RDW | 14.86 | 1.71 | % | 3.46% | 7.72% |
| WBC | 9.00 | 4.66 | 10^3/µL | 21.21% | 61.92% |

### **Table S6: Summary statistics of lab values.** These show the means and standard deviations (SDs) of the initial lab values and the percentage changes from these initial values for the data we analyze using the Markov model. Note, these summaries correspond to a different dataset (that obtained as described in the “Data processing for logistic and Markov models” section) than those provided in Table S1.

| Parameter(s) | Prior(s) | Reasoning |
| --- | --- | --- |
| Discharge individual patient intercepts and related population-level summaries | $\alpha_{0i}\sim normal\left( \alpha_{0}^{top}, \sigma_{\alpha} \right),$  $\alpha_{0}^{top}\sim normal\left( 0,1 \right),$  $\sigma_{\alpha}\sim normal\left( 0,0.05 \right)$ | Allows individual variation if substantial evidence exists |
| Expiry individual patient intercepts and related population-level summaries | $\beta_{0i}\sim normal\left( \beta_{0}^{top}, \sigma_{\beta} \right),$  $\beta_{0}^{top}\sim normal\left( 0,1 \right),$  $\sigma_{\beta}\sim normal\left( 0,0.05 \right)$ | Allows individual variation if substantial evidence exists |
| Regression coefficients | $\alpha_{1}\sim double\_exponential(0,1)$  $\beta_{1}\sim double\_exponential(0,1)$ | Sparsity inducing |

**Table S7: priors for Markov model parameters.**

### Data cleaning

We now detail the steps taken to clean and process the data, to convert it into a form amenable to estimation by both the logistic and Markov models.

There were n=926 clinical measurements where the test was recorded to take place before the date when the patient was admitted: in these instances, if the difference between date-time of admission and date-time of measurements was less than 48 hours (n=380 obs), we changed the date-time of admission for those individuals to be date-time of their first recorded test satisfying the 48 hour constraint; if the difference between date-time of admission and date-time of measurements exceeded 48 hours (n=546), the observation was dropped. Similarly, there were instances when measurements were reported after their recorded date-time of change of status (when either discharge or death occurred): if the gap between the date-time of their change of status and the observation exceeded 48 hours, the observation was dropped (n=274 obs); if this gap was less than 48 hours (n=68 obs), the date of change of status was changed to the latest observation within the 48 hour constraint. For many patients, there was a gap between their last observation and the date-time of their change of status. For those patients where this gap was less than a day, we updated the change of status date-time to the date-time of their last test (n=302 patients); for those patients who were eventually discharged but where the gap exceeded a day, we changed their change of status date-time to the date-time of their last test (n=134 patients). For the analyses involving time-dependent measurements, we dropped those patients who eventually died, where the gap between the date of the last observation and their date of death exceeded 48 hours (n=20 patients) as this typically signaled that the patients or their relatives had opted for palliative care. We dropped those patients where either the date-time of admission or the date-time of the change of status were unknown (n=6 patients).

### Data processing for logistic and Markov models

For the analyses involving time-dependent measurements, we carried out an additional series of transformations on the data. We then converted each patient's data into a regular day-block form: where, for each day the patient was in hospital, the patient had a single observation for each clinical test. To do this, we made a number of assumptions: if multiple tests of the same type were conducted on a patient on a given day, we took their mean as the daily observation; where a patient had a day when a specific test was not conducted, we assumed that day's test measurement was the same as on the last day it was conducted; when a patient was missing observations for a test until some days after their admission, we assumed the test measurements on these intervening days were the same as that from the first measurement. To reduce the impact of imputed observations, we included only those tests in the analysis where at least 70% of the individuals had a test taken (at least once); we then kept only those individuals who had been tested on at least 80% of the tests remaining after the previous step. Overall, approximately 27% of dynamic test observations were imputed, meaning that 27% of patient-test-days had observations imputed from previous or subsequent days' observations for that patient a described above. Finally, any individuals where age or sex were missing were dropped from the analysis. Collectively, these steps meant that n=475 patients with data from n=28 distinct clinical tests were included in the dynamic analyses.

For the analyses, we partitioned the influence of each clinical test into two separate variables: the initial measurement for each patient ("initial" meaning that the test was done within the first day of admission); and the percentage change in test value from this initial value. For those instances when an initial observation was zero, we set the percentage change as zero if all subsequent test values were also zero; alternatively, we calculated a percentage change versus half the first recorded non-zero value of the test. Finally, we standardized the percentage change data so that each test had a standard deviation of 1 and a mean of zero. Because of this, the effect sizes we estimate correspond to the typical scale of variation of each test (with those scales provided in Table S6).

### Inference models

#### Trend analysis

To compare trends in the clinical test values across the two patient groups (i.e. those that survived and those who died), we fit linear regression models incorporating time trends to the data. To do this, we used data for n=541 patients representing those remaining after removing those observations where either the date-time of admission or the date-time of the change of status were unknown and after having removed observations when the date-time of change of status occurred more than 48 hours after the date-time of the last observation.

For each combination of patient and test value, we calculated the percentage change in test value from the first test value taken on the patient (note that this calculation was slightly different to what was done for the logistic and Markov models). We then scaled this value by subtracting the mean and dividing through by the standard deviation in percentage test changes. Before conducting regressions, we also removed any infinite (i.e. when the first test value was zero) or missing observations for the percentage change. We also removed extreme observations: specifically, those where the absolute value of the percentage change exceeded the 98% quantile.

We modelled the percentage change in test value as a function of a quadratic time trend, allowing for fixed effect trends but including individual patient slopes of both the linear and quadratic terms of the trend. We fit separate models to each of the two patient groups and extracted the fixed effect estimates of the trends. The models were estimated in a frequentist framework using the lme4 R package (1).

#### Determining patient risk

We conducted two analyses of the data. The first was a logistic regression analysis, which aimed to determine those factors most predictive of patient mortality. The second was a Markov model, which analysed the dynamic sequence of observations for each patient throughout their stay and aimed to examine how changes in these variables affected how quickly a patient was discharged or died.

In the logistic regression, we examined how an individual patient's characteristics affected their risk of dying. This model is described by:

$y_{i}\sim\mathrm{Bernoulli}(\eta_{i}), \eta_{i}=\mathrm{logistic}(\delta_{0}+\boldsymbol{\delta}_{1}'\mathbf{x}_{i}),$

where $y_{i}\in\{0,1\}$ is the patient-specific outcome: $y_{i}=0$ represents discharge and $y_{i}=1$ represents death during the study period. The probability parameter of the Bernoulli model, $\eta_{i}\in[0,1]$, is given by a logistic transformation of a linear combination of the independent variables and parameters $\boldsymbol{\delta}_{1}$.

We performed five separate regression types, each with different groups of independent variables included. In the first of these, we included only a single variable in an analysis to examine the influence of each variable in isolation: so, this first analysis really consisted of a series of univariate logistic regressions: one for each of the variables included in the analysis. The second regression included patient characteristics not specifically relating to health ("patient" variables): their age, sex, ethnicity, and the day they were admitted to hospital. The third regression ("pat. + comorbidities") supplemented the background variables with the recorded comorbidities for each patient: whether they had hypertension, diabetes etc. (13 conditions in total). The fourth regression ("admission") supplemented the third with the initial measurements for each patient for each of the 28 included clinical tests. The final regression ("post-admission") then included the percentage changes in each clinical test measurement from the initial values for each patient: this was calculated using the last recorded measurement for each patient.

We next sought to model not only the outcome of each patient but also the time taken for this outcome to be reached. An approach that is often used to analyse these sorts of data in the literature is Cox regression incorporating competing risks. We reviewed these models and decided not to use these due to issues with the assumptions underpinning them: specifically, that those individuals who are discharged would effectively be handled as if they were uninformatively censored observations to determine a mortality risk. Instead, we chose to develop a Markov model that simultaneously modelled individuals who were discharged and those who died. To do so, we considered the sequence of outcomes for each day the patient remained in hospital. On their first day, they were admitted and began in the "hospital" state (see Fig. 1); at the end of the first day, they either remained in hospital or transitioned to the "discharge" or "death" states. On subsequent days, the patients that remained in hospital faced the same possible transitions. The probabilities that these transitions occurred were modelled as a function of each patient's characteristics: some of which could potentially vary over time. Specifically, the un-normalised probabilities of each possible transition were modelled using a log link:

$$q_{it}^{\mathrm{discharge}}=\exp(\alpha_{0i}+\boldsymbol{\alpha}_{1}'\mathbf{x}_{it}), q_{it}^{\mathrm{death}}=\exp(\beta_{0i}+\boldsymbol{\beta}_{1}'\mathbf{x}_{it}), q_{it}^{\mathrm{hospital}}=1,$$

where $i$ indicates a patient id; $t$ represents the day being considered; $\boldsymbol{\alpha}_{1}$and $\boldsymbol{\beta}_{1}$ are vectors of regression coefficients; $\mathbf{x}_{it}$ is a vector of regressors (some of which may be time dependent); and $\alpha_{0i}$ and $\beta_{0i}$ are time-constant patient-specific parameters. To compute normalised probabilities, we used a softmax transformation:

$$p_{it}^{\mathrm{discharge}}=q_{it}^{\mathrm{discharge}}/q_{it}^{\mathrm{total}}, p_{it}^{\mathrm{death}}=q_{it}^{\mathrm{death}}/q_{it}^{\mathrm{total}}, p_{it}^{\mathrm{hospital}}=q_{it}^{\mathrm{hospital}}/q_{it}^{\mathrm{total}},$$

where $q_{it}^{\mathrm{total}}=q_{it}^{\mathrm{discharge}}+q_{it}^{\mathrm{death}}+q_{it}^{\mathrm{hospital}}$. Here, $p_{it}^{\mathrm{discharge}}$, $p_{it}^{\mathrm{death}}$ and $p_{it}^{\mathrm{hospital}}$ represent the probabilities of being discharged, dying or remaining in hospital on a given day, respectively.

Due to computational expense, we did not calculate univariate odds ratios. As for the logistic regression case, we considered the same four sets of multivariate regressors. The only difference was that, for the post-admission case, the percentage change variables represented those values on each specific day $t$, rather than those values at the end of a patient's stay in hospital.

Both analyses were conducted in a Bayesian framework. In both cases, we used priors for the regression coefficients $(\boldsymbol{\delta}_{\mathbf{1}},\boldsymbol{\alpha}_{\mathbf{1}},\boldsymbol{\beta}_{\mathbf{1}})$ meant due induce sparsity in the estimates: meaning that only those most significant factors would be estimated to have non-zero effects. For the logistic model, we used "horseshoe" priors (2); for the Markov model, we used Laplace priors instead since we had convergence issues when attempting to use the horseshoe. For the Markov model, the priors used are shown in Table S7.

The models were fit using Markov chain Monte Carlo, and we used Stan's dynamic Hamiltonian Monte Carlo (3) for sampling. For the logistic model, the models were written using the rstanarm package (4) and were run with 2000 iterations across each of 4 Markov chains. For the Markov model, the models were written using Stan using the following code:

data{

int N; // num obs

int patient[N]; // indicates patient identification

int npatient; // num patients

int state[N]; // either 1 (discharge), 2 (in hospital) or 3 (death) during each day

int ncovs; // number of covariates included in regression model

matrix[N, ncovs] X; // regressor matrix

}

transformed data{

vector[3] ones = to_vector(rep_array(1, 3));

matrix[N, 3] mstate;

for(i in 1:N)

for(j in 1:3)

mstate[i, j] = state[i] == j ? 1 : 0;

}

parameters{

vector[npatient] a0_raw;

real a0_top;

real<lower=0> sigma_a0;

vector[npatient] b0_raw;

real b0_top;

real<lower=0> sigma_b0;

vector[ncovs] a1;

vector[ncovs] b1;

}

transformed parameters {

// non-centered parameterisation for intercept priors

vector[npatient] b0 = b0_top + sigma_b0 * b0_raw;

vector[npatient] a0 = a0_top + sigma_a0 * a0_raw;

}

model{

matrix[N, 3] p;

vector[N] psum;

p[, 1] = exp(a0[patient] + X * a1);

p[, 3] = exp(b0[patient] + X * b1);

p[, 2] = to_vector(rep_array(1, N));

psum = p * ones; // sums each row

p = p ./ rep_matrix(psum, 3); // normalises each row of probability matrix

p = p .* mstate; // zeros any probs corresponding to states that weren’t observed

target += log(p * ones); // likelihood

// priors

b0_raw ~ normal(0, 1);

b0_top ~ normal(0, 1);

sigma_b0 ~ normal(0, 0.05);

a0_raw ~ normal(0, 1);

a0_top ~ normal(0, 1);

sigma_a0 ~ normal(0, 0.05);

b1 ~ double_exponential(0, 1);

a1 ~ double_exponential(0, 1);

}

The Markov models were run using 4000 iterations although these were thinned by a factor of 2 for space. For both analyses, the first half of the iterations were discarded as warm-up, and $\hat{R}<1.01$ for all parameters providing evidence of convergence (5).

The results produced for each analysis depend on the model providing a reasonable approximation to the actual data generating process. To check this assumption, for each analysis, we performed a series of posterior predictive checks, which compare the actual data with that simulated from the model (6). These graphical checks are shown in Figs. S6-S9, and indicated that the logistic and Markov models including all the available independent variables in it (the *post-admission* regression) provided a good fit to the data.

#### Markov model checking using cross-validation

As per PROBAST guidelines (7), in lieu of not having access to alternative data, we tested the performance of our model by splitting our data into training and testing sets. We used an approach known as K-fold cross-validation, where the dataset is repeatedly split into training and testing sets, such that each observation appears only once in each of the testing sets. Specifically, we used 5 folds, where each testing set was of size 95 and generated by selecting observations uniformly at random from the overall set (without replacement). For each fold, we fitted each of the four Markov models on the training set using MCMC (via Stan) with 500 post-warm-up draws (and 500 pre-warm-up draws) per chain across 4 chains. We then used the trained model to predict the outcome on the independent testing set. For each individual, we generated a predicted outcome for each MCMC draw, meaning that we could, in principle, have a range of outcomes across the entire MCMC sample. To compare the predicted outcome with the actual, we took the median prediction and use that to calculate a prediction accuracy on each testing set. Examining the variation in accuracy across all the testing sets then yields a measure of uncertainty.
